# Antiseizure medication withdrawal risk estimation and recommendations: a survey of American Academy of Neurology and EpiCARE members

**DOI:** 10.1101/2022.11.29.22282905

**Authors:** Samuel W Terman, Renate van Griethuysen, Carole E Rheaume, Geertruida Slinger, Anisa S Haque, Shawna N Smith, Wesley T Kerr, Charlotte van Asch, Willem M Otte, Carolina Ferreira-Atuesta, Marian Galovic, James F Burke, Kees PJ Braun

**Author notes:** Corresponding author: Samuel W Terman, MD MS, University of Michigan Department of Neurology, Department of Neurology, Taubman 1^st^ Floor, Reception C, 1500 E Medical Center Dr, SPC, 5316. Ann Arbor, MI 48109. Additional author email addresses: Ms Griethuysen Ms Rheaume Dr Slinger Ms Haque Dr Smith Dr Kerr Dr van Asch Dr Otte Dr Ferreira-Atuesta Dr Galovic Dr Burke Dr Braun.

## Abstract

**Background:** Choosing candidates for antiseizure medication (ASM) withdrawal in well-controlled epilepsy is challenging. We evaluated 1) the correlation between neurologists’ seizure risk estimation (“clinician predictions”) versus calculated predictions, 2) how viewing calculated predictions influenced recommendations, and 3) barriers to using risk calculation.

**Methods:** We asked neurologists to predict two-year seizure risk after ASM withdrawal for hypothetical vignettes. We compared withdrawal recommendations ASMs before versus after viewing calculated predictions using generalized linear models.

**Results:** Three-hundred forty-six responded. There was moderate correlation between clinician and calculated predictions (Spearman coefficient 0.42). Clinician predictions varied widely, e.g., predictions ranged 5%-100% for a two-year seizure-free adult without epileptiform abnormalities. Mean clinician predictions exceeded calculated predictions for vignettes with epileptiform abnormalities (e.g., childhood absence epilepsy: clinician 65%, 95% confidence interval [CI] 57%-74%; calculated 46%) and surgical vignettes (e.g., focal cortical dysplasia six-months seizure-free mean clinician 56%, 95% CI 52%-60%; calculated 28%). Clinicians overestimated the influence of epileptiform EEG findings on withdrawal risk (26%, 95% CI 24%-28%) compared with calculators (14%, 95% 13%-14%). Viewing calculated predictions slightly reduced willingness to withdraw (−0.8/10 change, 95% CI -1.0 to -0.7), particularly without epileptiform abnormalities. The greatest barrier to calculator use was doubting its accuracy (44%).

**Conclusions:** Clinicians overestimated the influence of abnormal EEGs particularly for low-risk patients and overestimated risk and the influence of seizure-free duration for surgical patients, compared with calculators. These data may question widespread ordering of EEGs or time-based seizure-free thresholds for surgical patients. Viewing calculated predictions reduced willingness to withdraw particularly without epileptiform abnormalities.

**What is already known on this topic:** For the two-thirds of patients with epilepsy who become seizure-free on antiseizure medications (ASMs), a key question is whether ASMs are necessary indefinitely after attaining seizure-freedom. An individualized post-withdrawal seizure risk calculator exists,[1,2] which has demonstrated moderate external validity, and one 1993 study suggested that viewing results from an older version of such a calculator slightly reduced patients’ willingness to withdraw.

**What this study adds:** Before our study, it was unknown how closely clinicians’ intuitive estimates of post-withdrawal seizure risk (“clinician predictions”) align with model (“calculated”) predictions, and no study had previously evaluated the influence of viewing calculated risk on clinician recommendations. We found 1) moderate correlation between clinician and calculated predictions (Spearman coefficient 0.42), 2) clinician predictions and recommendations varied widely, 3) clinicians tended to overestimate risk compared with calculators for vignettes with epileptiform abnormalities and surgical cases, 4) viewing calculated predictions slightly reduced willingness to withdraw particularly without epileptiform abnormalities, and 5) the greatest endorsed barrier to calculator use was doubting its accuracy.

**How this study might affect research, practice or policy:** Our study suggests patient scenarios in which viewing calculator results may be most useful or be most likely to change clinical practice. These data may also question widespread ordering of EEGs or time-based seizure-free thresholds for surgical patients given calculated risk differences were smaller than clinician predictions. We also provide preliminary data guiding future work to improve calculator implementation such as targeting educational efforts to improve knowledge of its development, improving its accuracy, and integrating it into the electronic medical record.

## Introduction

Two-thirds of patients with epilepsy become seizure-free on antiseizure medications (ASMs).[3] For these patients, a key question is whether ASMs are necessary indefinitely. ASMs reduce morbidity and improve quality of life by decreasing seizures.[4,5] However, adverse effects reduce quality of life,[6–9] and risk declines with longer seizure-freedom.[10,11] Thus, guidelines have endorsed considering withdrawal after detailed counseling.[12–14]

Accurate post-withdrawal seizure risk is key to optimizing decision-making. Physicians often overestimate treatment benefits and underestimate harms, and risk calculators tend to outperform humans at estimating risk.[15] Individualized post-withdrawal seizure risk calculators exist for medical[1,2] and surgical[16] patients. Yet, the degree to which clinicians’ intuitive estimate of post-withdrawal seizure risk (“clinician predictions”) align with model (“calculated”) predictions remains unclear, which may inform when viewing calculated results might most useful. Furthermore, recent guidelines highlight the need for data evaluating the influence of post-ASM withdrawal prediction tools on recommendations.[14] One study found that viewing calculated predictions decreased willingness to withdraw ASMs.[17] However, that included a small single-center sample, did not include children or post-surgical cases, and was based on a now-outdated calculator, thus applicability to current practice is unclear. Another survey documented variability in clinicians’ ASM withdrawal recommendations, but did not assess clinicians’ risk predictions or the influence of viewing calculator results.[18]

To better understand current physician practice patterns surrounding ASM withdrawal decisions, we conducted an international survey of neurologists. We evaluated 1) the correlation between clinician predictions and calculated predictions, 2) how viewing calculated predictions influences ASM withdrawal recommendations, and 3) barriers to using risk calculation.

## Methods

### Participants

We recruited neurologists. This included epileptologists and non-epileptologists, to compare responses, and given many non-epileptologists care for epilepsy patients.[19] The first group included current US AAN members. Invitees were ≤68 years old, practicing in general neurology or with a subspecialty of epilepsy or clinical neurophysiology, and had not responded to another AAN survey in the previous 6 months. This left 5,649 US AAN members; we randomly sampled 4,001 (to leave participants for other AAN survey priorities). The second group consisted of all 403 eligible European AAN members, and a third non-AAN group included 519 EpiCARE members. EpiCARE is a European network treating complex epilepsies, spanning 28 institutions across 24 countries. We asked respondents to disregarding duplicate invitations if they were European AAN and EpiCARE members. We kept only the first attempt for 13 respondents who began the survey twice. Individuals received up to three email reminders, with recruitment spanning five months. There was no compensation.

### Procedures involving human subjects

This study was approved by the University of Michigan IRB. The first page of the survey requested informed consent.

### Survey variables and design

We assembled experts in ASM withdrawal decisions and developed a cross-sectional survey collaborating with the American Academy of Neurology (AAN) (Appendix). The research team met to discuss objectives and drafted content to address objectives. All members (including an implementation scientist, epileptologists, non-epileptologist neurologists, statisticians, and a research coordinator) pretested the survey, and questions were refined until all members agreed that questions were clear and met objectives. The survey was designed in English, programmed into Qualtrics, and distributed electronically.

To evaluate for nonresponse bias, we compared demographics of all invited AAN members stratified by whether they consented to take the survey. These data included age, sex, race, geographic region, academic versus non-academic practice, percent of time spent in research, and subspecialty. This was possible only for AAN members, because AAN’s member database contains demographic information for all members, whereas EpiCARE data was available only from respondents.

We asked respondents whether they treat mostly adults or children, what percent of their patient practice is spent evaluating seizures or patients with epilepsy surgery, whether they are board-certified in epilepsy or clinical neurophysiology, and their years of experience treating epilepsy patients.

While it is impossible to design vignettes presenting the full complexity of clinical care, we developed specific vignettes intended to represent a broad range of representative cases (Supplemental Table 1). Vignettes spanned children and adults, surgical and non-surgical (“medical”) cases, different epilepsy etiologies, and different durations of seizure-freedom.

Respondents were randomized to complete half of the clinical scenarios for which they were eligible, based on whether they treat adults versus children, and any post-surgical patients. Respondents viewed medical then surgical vignettes, if they treat both populations. We did not present pediatric surgical vignettes, given a previous overlapping study.[20]

Each vignette began with a reported normal EEG (we did not show actual EEG images). We asked respondents to estimate the patient’s risk of having another seizure in the next two years (“clinician prediction”) if the patient withdrew versus continued ASMs. We did not specify any precise withdrawal schedule or duration, given no significant between faster versus slower tapering.[14,21] We also asked how likely the respondent would be to recommend withdrawal on a Likert scale (0/10: extremely unlikely; 5/10: neither likely nor unlikely; 10/10: extremely likely) under different scenarios such as whether the patient’s job required driving, whether they were experiencing ASM-related side effects, or wished for future pregnancy. Then, we repeated these questions in the presence of interictal epileptiform EEG abnormalities.

For medical vignettes, we then showed respondents calculated predictions of two-year seizure relapse risk if the patient withdrew versus continued ASMs and requested an updated recommendation regarding how likely they would be to advise withdrawal. We obtained post-withdrawal calculated predictions from Lamberink and colleagues’ online risk calculator (http://epilepsypredictiontools.info/aedwithdrawal). This calculator was developed from 1,769 patients pooling 10 real-world datasets, demonstrated moderate performance during internal-external cross-validation (area under the curve 0.65), and provides the most rigorous currently available individualized post-withdrawal seizure risk prediction.[1] To obtain ‘continuation’ calculated predictions of two-year seizure relapse, we multiplied post-withdrawal calculated predictions by 50%, as informed by two randomized trials. 1) The largest randomized trial to date found 41% versus 22% relapse by two years.[22] 2) The other trial relevant to adults found a one-year relapse risk of 15% versus 7% during double-blinded follow-up.[23] No literature currently enables further individualized relative risk reductions.

For surgical vignettes, we similarly obtained clinician predictions and withdrawal recommendations. However, while a surgical post-withdrawal online risk calculator has very recently been validated (Ferreira-Atuesta and colleagues[16]; https://predictepilepsy.github.io/), its validation was not yet complete at the time of this survey. Thus, we incorporated surgical calculated predictions into our analysis but not the actual survey. We also did not incorporate post-surgical continuation calculated risks, in absence of RCT data.

### Statistical analysis

We compared clinician versus calculated predictions using scatterplots and Spearman’s correlation coefficients with 1,000 bootstrapped replications to obtain confidence intervals [CI].

We also displayed violin plots of clinician versus calculated predictions by vignette. Because Lamberink model overpredicted risk in two external validation studies,[24,25] and a third validation study demonstrated poor calibration,[26] we performed a sensitivity analysis using external predictions; we chose the predicted risk from the Lamberink model that corresponded to the published logistic calibration curve from Lin and colleagues (their Figure 4a).[24] To compute mean clinician predictions with 95% CI’s for each vignette and thus to compare mean clinician versus calculated predictions, we then performed generalized linear models.[27] The outcome consisted of clinician predictions (0%-100%) for each vignette. Covariates included vignette, ASM withdrawal versus continuation, epileptiform EEG, and all their pairwise interactions. We used a logit link, binomial family, and cluster robust standard errors for respondents. Note that the Ferreira-Atuesta surgical calculator did not include EEG results because some centers contributing data to the model did not have EEG information. Their model also contained one variable that we did not specify in our vignettes (presurgical seizure frequency), because that variable was added to the model after our survey data collection was already in progress. Therefore, in our analyses for surgical vignettes we averaged across EEG results and displayed sensitivity results for Ferreira-Atuesta results assuming either monthly or weekly presurgical seizure frequency.

We performed secondary analyses regarding differences between clinician versus calculated predictions. We first assessed more broadly how vignette-related and respondent-related covariates influenced differences using generalized linear models. The outcome was the difference between clinician minus Lamberink calculated predictions, with a Gaussian link function. We repeated this model using calculated predictions from the Lin model. Additionally, to address the survey’s low response rate, we used inverse probability of selection weighting.[28] Inverse probability of selection weighting seeks to mitigate selection bias by upweighting respondents who had a low predicted probability of participating, to simulate a dataset as if all participants had responded. To do so, first we performed a logistic regression for whether invited AAN members consented to participate. We used independent variables that might influence participation: age, sex, race, region of the US, academic versus nonacademic practice, percent time clinical versus research, and epileptologist/clinical neurophysiologist. Then, we repeated the main analysis, weighting each respondent by one divided by the predicted probability of consenting to participate.

We then assessed whether viewing calculated predictions changed recommendations (“pre” versus “post” viewing calculated predictions) using scatterplots, violin plots, and bar charts. Recommendations took on values between 0 (extremely unlikely to recommend withdrawal) and 10 (extremely likely). We divided responses by 10 to bound the outcome between 0 and 1 to facilitate using generalized linear models with a logit link. Each recommendation was an outcome, whether the vignette displayed calculated risks was a covariate, and we used cluster-robust standard errors to account for within-respondent correlation. Sensitivity models performed inverse probability of selection weighting. We also performed secondary analyses showing scatterplots and lowess curves to assess the degree to which recommendations correlated with risk, and then displayed all results from our generalized linear model describing what respondent and vignette factors influenced recommendations.

### Data availability statement

The American Academy of Neurology is the sole owner of the raw data.

## Results

### Sample characteristics

We sent the survey to 4,923 individuals, of whom 463 consented, 411 passed eligibility questions, and 346 responded to at least one vignette. AAN members consenting to participate were more likely than those not consenting to be White, European, academic, specialized, and researchers (Table 1).

**Table 1:** Population description. To evaluate for selection bias, we compared AAN member survey recipients who did versus did not consent to complete the survey (“AAN data only”). P-values compare AAN members who did versus did not consent using Chi-squared and t-tests. For respondents, we then displayed information combining AAN member data plus information they provided during the survey itself to describe our included population.

|  |  | Did not consent | Consented | p | Include |
| --- | --- | --- | --- | --- | --- |
| From AAN data only, to assess selection into the study |  |  |  |  |  |
| N |  | 4,036 (92%) | 368 (8%) |  | 346* |
| Age, years |  | 50 (41-58) | 51 (42-60) | 0.14 | 50 (42-60) |
| Female |  | 1,513/3,957 (38%) | 147/364 (40%) | 0.42 | 144/332 (43%) |
| Race | White | 1,943/3,304 (59%) | 220/329 (67%) | <0.01 | 178/255 (70%) |
|  | Black | 87/3,304 (3%) | 14/329 (4%) |  | 11/255 (4%) |
|  | Asian | 805/3,304 (24%) | 56/329 (17%) |  | 38/255 (15%) |
| Region of US | Northeast | 812/3,663 (22%) | 63/311 (20%) | 0.69 | 44/240 (18%) |
|  | Midwest | 722/3,663 (20%) | 69/311 (22%) |  | 57/240 (24%) |
|  | South | 1,318/3,663 (36%) | 113/311 (36%) |  | 91/240 (38%) |
|  | West | 811/3,663 (22%) | 66/311 (21%) |  | 48/240 (20%) |
| Cohort | AAN – US | 3,687 (91%) | 314 (85%) | <0.01 | 242 (70%) |
|  | AAN – Europe | 349 (9%) | 54 (15%) |  | 46 (13%) |
|  | EpiCARE | - | - |  | 58 (17%) |
| Academic** |  | 785/3,993 (20%) | 111/367 (30%) | <0.01 | 91/287 (32%) |
| Percent clinical** |  | 90% (75%-100%) | 80% (60%-95%) | <0.01 | 80% (60%-95%) |
| Percent research |  | 0% (0%-5%) | 0% (0%-8%) | <0.01 | 0% (0%-8%) |
| Epilepsy/CNP |  | 2,043 (51%) | 240 (65%) | <0.01 | 186/288 (65%) |
| From all data, to describe the respondents |  |  |  |  |  |
| Academic** |  | - | - |  | 173 (50%) |
| Percent clinical** |  | - | - |  | 90% (75%-100%) |
| % of patients seen for seizures | <50% | - | - |  | 143 (42%) |
|  | ~50% | - | - |  | 45 (13%) |
|  | >50% | - | - |  | 155 (45%) |
| % of patients post-epilepsy surgery | None | - | - |  | 115 (33%) |
|  | Some | - | - |  | 229 (66%) |
|  | All | - | - |  | 2 (1%) |
| Population | Mostly ≥18 yrs | - | - |  | 268 (77%) |
| Board-certification | Epilepsy | - | - |  | 143/277 (52%) |
|  | CNP | - | - |  | 120/277 (43%) |
| Specialty | Epilepsy |  | - |  | 209/335 (62%) |
|  | CNP |  | - |  | 158/335 (47%) |
| Years treating patients with epilepsy | 0-5 | - | - |  | 33/277 (12%) |
|  | 6-10 | - | - |  | 47/277 (17%) |
|  | 11-15 | - | - |  | 41/277 (15%) |
|  | 16-20 | - | - |  | 31/277 (11%) |
|  | 20+ | - | - |  | 125/277 (45%) |

AAN: American Academy of Neurology; CNP: clinical neurophysiology.
\*These 346 respondents included 242 US AAN members (4,001 sent, 314 consented, 287 eligible, 197 completed), 46 European AAN members (403 sent, 54 consented, 49 eligible, 46 completed), and 58 EpiCARE members (519 sent, 95 consented, 75 eligible, 58 completed). Countries represented included the US (242), Germany (11), Italy (11), Switzerland (8), United Kingdom (7), Portugal (6). The remaining all had less than 5: Austria, Belgium, Croatia, Cyprus, Czech Republic, Denmark, Finland, France, Georgia, Hungary, Ireland, Latvia, Lithuania, Luxembourg, Netherlands, Poland, Romania, Russia, Serbia, Slovenia, Spain, Sweden.
\*\*Note variables for academic practice and percent clinical are repeated in the table. The first mention is based only on variables contained in the database of AAN member information that AAN members reported at the time of creating their AAN profile. The second mention of 'academic' is whether the individual reported being in an academic practice either through their AAN profile, or else at the time of taking our survey (and thus this variable is reported only for the 'included' respondents, and hence higher than from AAN profile data alone. The second mention of 'percent clinical' is based on responses at the time of taking our survey.

### Vignette responses: risk estimation

Clinician risk predictions were highly variable across vignettes, often ranging from nearly 0% to 100% (Figure 1; Supplemental Figure 1). For example, predictions ranged from 5% to 100% for a two-year seizure-free adult without epileptiform abnormalities (5^th^ percentile: 19%; 25^th^ percentile: 34%; 50^th^ percentile: 50%; 75^th^ percentile: 62%; 95^th^ percentile: 86%). Fifty-eight percent of clinician predictions were within 20% of calculated predictions. Clinician and calculated predictions demonstrated moderate correlation (0.42, 95% CI 38%-46%).

**Figure 1.**
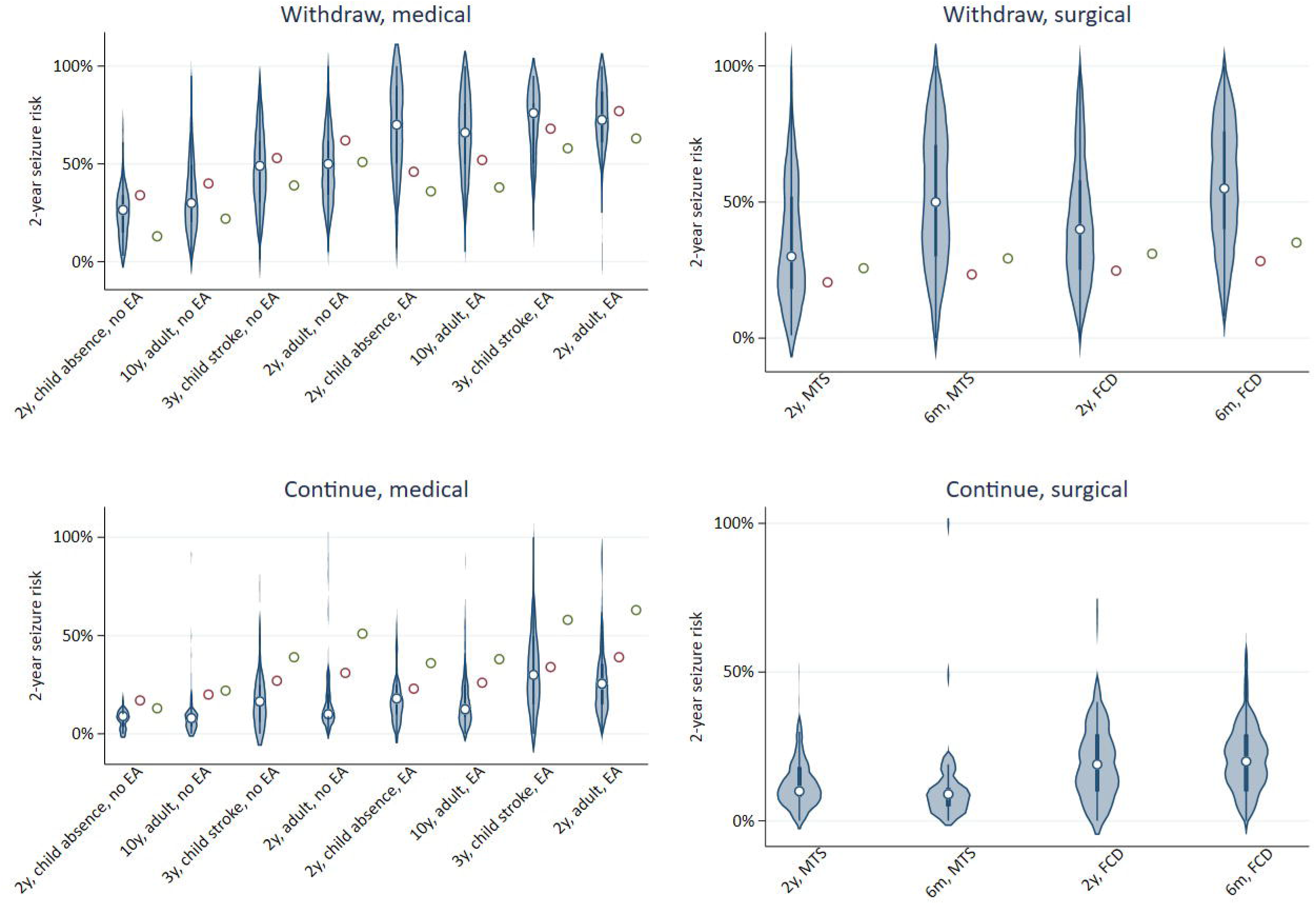
Distribution of clinician versus calculated predictions of two-year seizure relapse for each vignette. Left: medical vignettes; Right: surgical vignettes; Top: withdraw antiseizure medication (ASM); Bottom: continue ASM. Medical vignettes are further separated according to epileptiform EEG abnormalities (left half of medical panels) and no epileptiform EEG abnormalities (right half of medical panels). For medical panels, circles represent calculated predictions from Lamberink (red) or Lin (green) models. For the surgical withdrawal panel, circles represent calculated predictions from Ferreira-Atuesta assuming a monthly (red) or weekly (green) presurgical seizure frequency. *Interpretation*: There was wide variation in clinical versus calculated predictions of seizure risks throughout vignettes. MTS: mesial temporal sclerosis; FCD: focal cortical dysplasia.

Mean clinician predictions for medical vignettes were almost all within 10%-20% of calculated predictions (Figure 2). Clinician predictions for post-withdrawal medical vignettes most exceeded calculated predictions for vignettes with epileptiform EEG findings – (1) the two-year seizure-free child with absence epilepsy (mean clinician prediction 65%, 95% CI 57%-74%; Lamberink calculated prediction 46%, Lin calculated prediction 36%; p<0.05), and (2) the ten-year seizure-free adult (mean clinician prediction 64%, 95% CI 60%-68%; calculated Lamberink prediction 52%, calculated Lin prediction 38%; p<0.05). Clinician predictions tended to be lower than Lamberink calculated predictions across medical vignettes without epileptiform EEG findings (p<0.05 for each) and most continuation vignettes.

**Figure 2.**
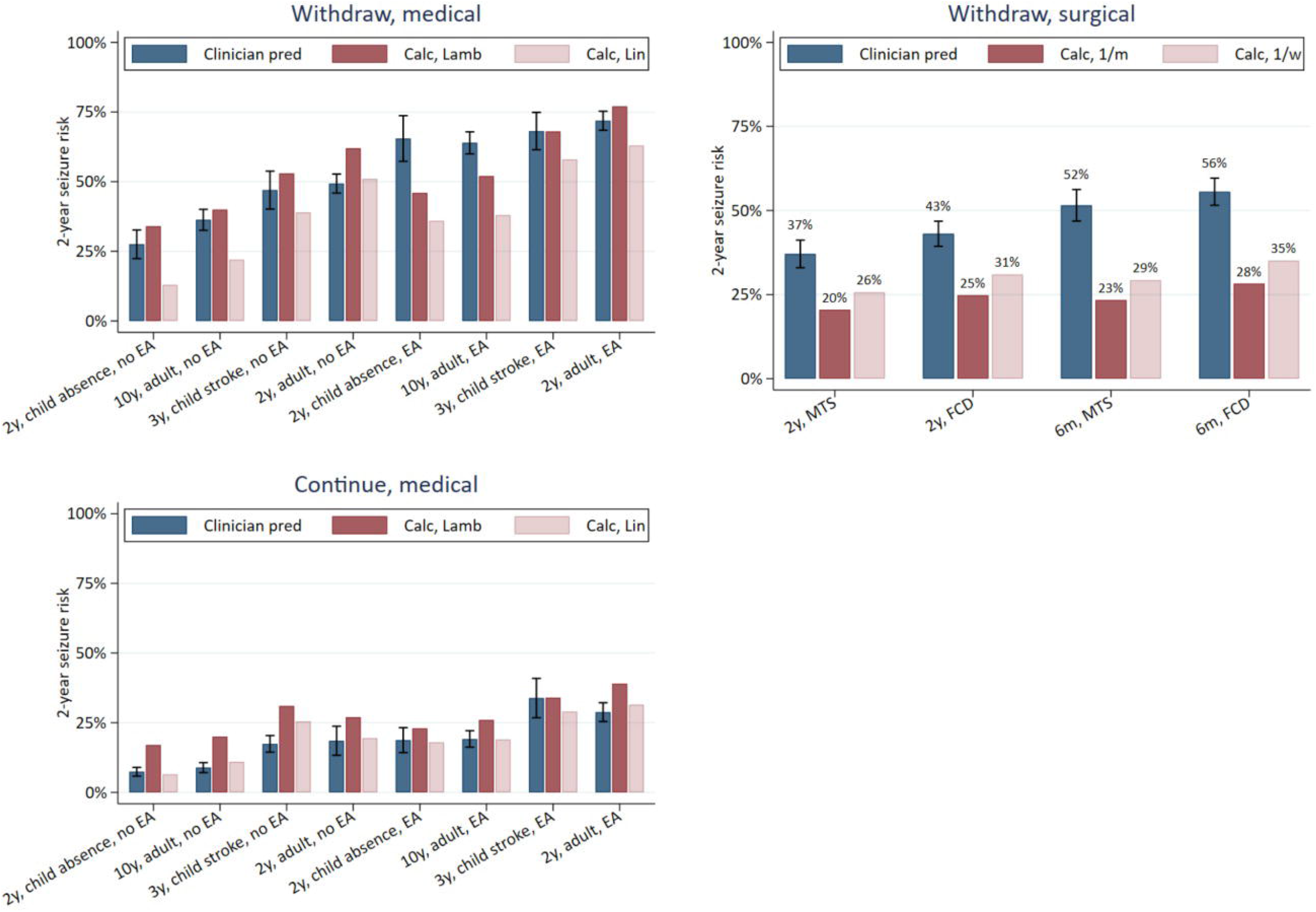
Mean and 95% confidence intervals for clinician versus calculated predictions of two-year seizure relapse risk by vignette. *Interpretation:* Mean clinician predictions for medical vignettes were mostly within 10%-20% of calculated risks. Clinician predictions were particularly higher than calculated predictions for post-withdrawal vignettes with epileptiform EEG findings for the two-year seizure-free child with absence epilepsy and the ten-year seizure-free adult. Clinician predictions were higher than calculated predictions for all surgical vignettes, but lower than calculated predictions for continuation medical vignettes without epileptiform EEG abnormalities. Note there is no “Continue, surgical” panel as there was in Figure 1 because randomized data does not yet exist enabling a calculated continuation surgical risk to compare with responses.

Clinician predictions for post-withdrawal surgical vignettes all exceeded calculated predictions (e.g., focal cortical dysplasia six-months seizure-free mean clinician prediction 56%, 95% CI 52%-60%; calculated prediction 28%; p<0.05).

Clinicians estimated a greater influence of epileptiform EEGs on post-withdrawal risk (26%, 95% CI 24%-28%) compared with calculated predictions (14%, 95% CI 13%-14%). Respondents who indicated they always order EEGs before considering withdrawal estimated an epileptiform EEG to confer greater risk than respondents who do not always order EEGs (Supplemental Figure 2).

Mean clinician predictions were generally similar across vignettes when comparing epileptologists/clinical neurophysiologists versus all other respondents (Supplemental Figure 3, which recapitulates Figure 2 except including stratification by specialization). Most respondent characteristics only slightly influenced adjusted differences between clinician and calculated predictions (Supplemental Figure 4). An exception was that respondents who see only surgical epilepsy patients provided lower clinician predictions, though the confidence interval was wide given only N=16 (compared with 2,020 predictions for respondents who see a mixture of medical/surgical patients and 450 predictions for respondents who only see medical patients). Rather, differences were driven by vignette characteristics. Factors associated with higher clinician than calculated predictions included surgical cases, pediatric cases, longer seizure-free periods, epileptiform EEG, and post-withdrawal cases (p<0.05). Inverse probability of selection weighting yielded similar conclusions (Supplemental Figure 5).

### Vignette responses: recommendations

Viewing calculator results made respondents less likely to recommend withdrawal (Figure 3; pre: mean 3.5/10; post: 2.7/10; mean change -0.8, 95% CI -1.0 to -0.7; p<0.05), particularly for respondents most likely to recommend withdrawal at baseline. Variation in recommendations was wide for all vignettes (Figure 4), often ranging between 0/10 and 10/10. Respondents were least likely to recommend withdrawal for a two-year seizure-free adult with epileptiform EEG abnormalities (mean recommendation before viewing calculator: 1.3/10; mean recommendation after viewing calculator: 1.2/10). Respondents were still unlikely to recommend withdrawal for the two-year seizure-free adult even with a normal EEG (before seeing risk: 3.3; after: 1.9/10), and viewing the calculator most reduced recommendations in cases with a normal EEG (Figure 5). Respondents were the most likely to recommend withdrawal for a child with absence epilepsy and a normal EEG.

**Figure 3.**
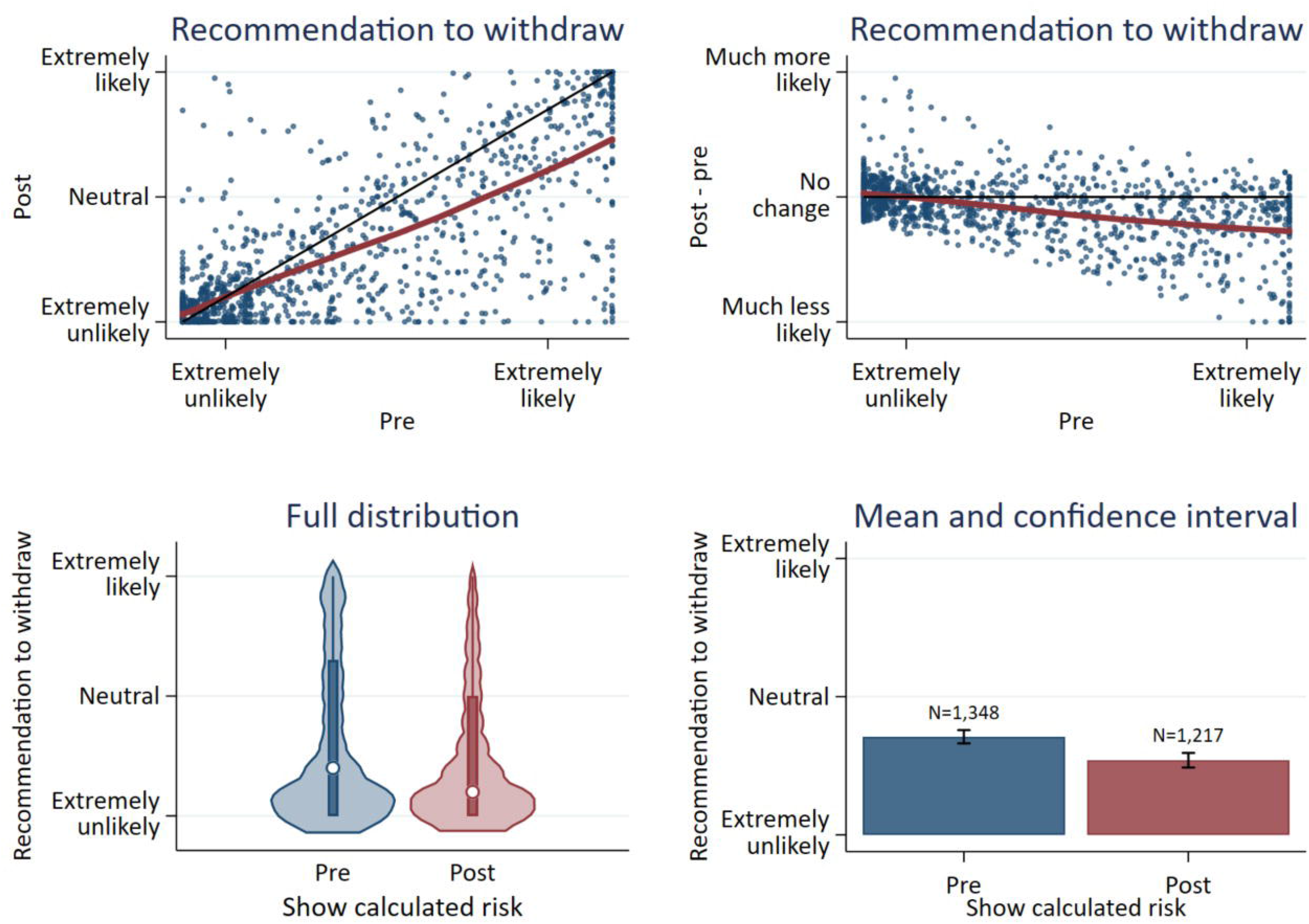
Likelihood to recommend withdrawal before (“pre”) versus after (“post”) viewing results from the calculator for medical vignettes. Upper panels represent scatterplots comparing pre- versus post-recommendations (left) or comparing how much recommendations changed after viewing calculated predictions versus “pre” recommendations. Black lines would represent no change, and red curves are loess curves fit to the data. *Interpretation:* Viewing calculator results on average made respondents slightly less likely to recommend withdrawal, particularly for respondents who were more likely to recommend withdrawal at baseline.

**Figure 4.**
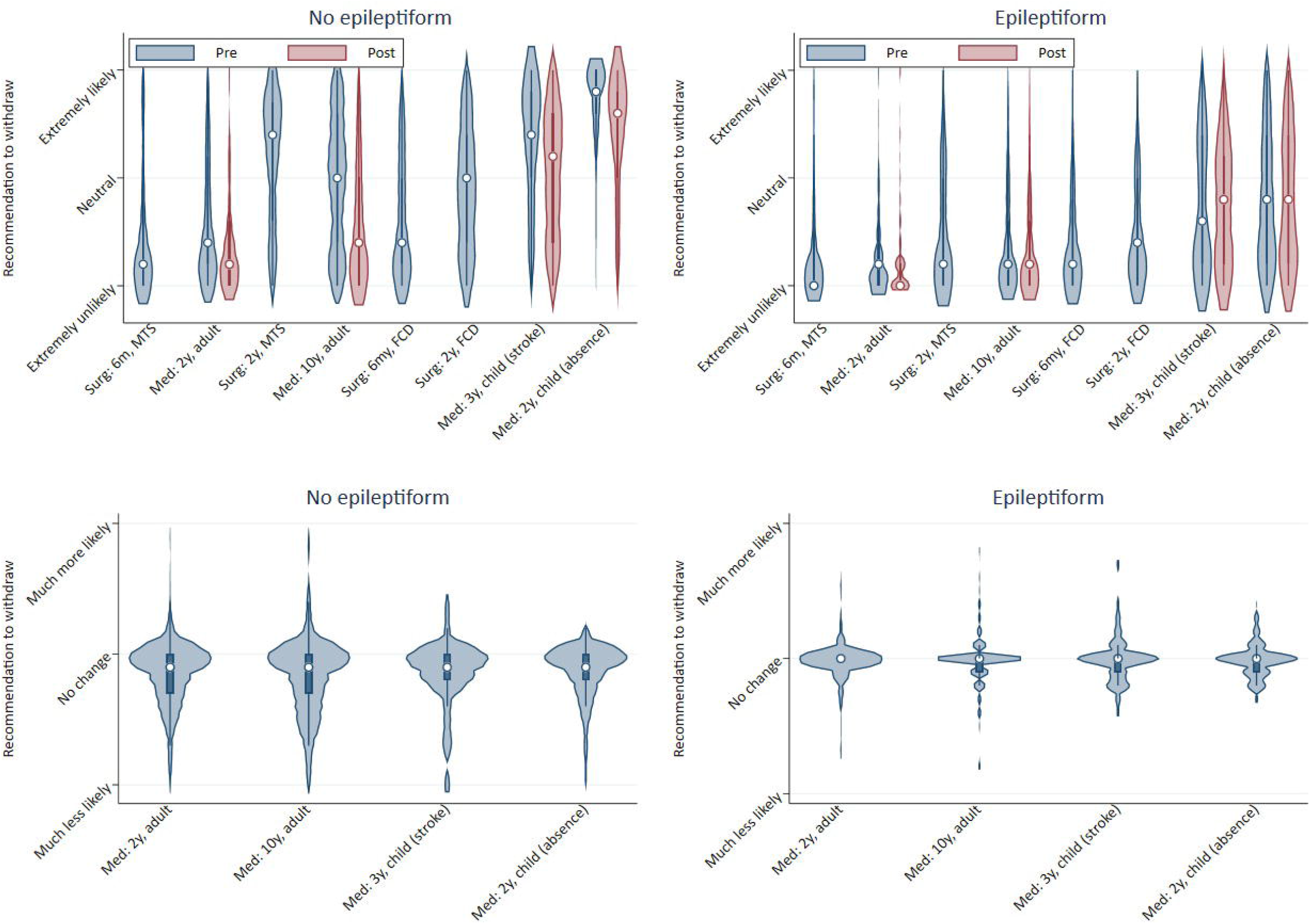
Recommendation to withdraw before (pre) versus after (post) viewing results of the calculator. Top: Distribution of recommendations for each vignette pre versus post viewing results of the calculator (only medical vignettes had a ‘post’ available). Bottom: Distribution of changes in recommendations, post minus pre. *Interpretation:* Variation in recommendations was wide for all vignettes, ranging in many vignettes between 0/10 (extremely unlikely) and 10/10 (extremely likely). On average, viewing calculator results made little difference in recommendations, or else slightly reduced recommendations to withdraw particularly in vignettes without epileptiform EEG abnormalities. Surg: surgical vignette; Med: medical vignette; 6m: 6 months seizure-free; 2y: 2 years seizure-free; 3y: 3 years seizure-free; 10y: 10 years seizure-free; MTS: mesial temporal sclerosis; FCD: focal cortical dysplasia.

**Figure 5.**
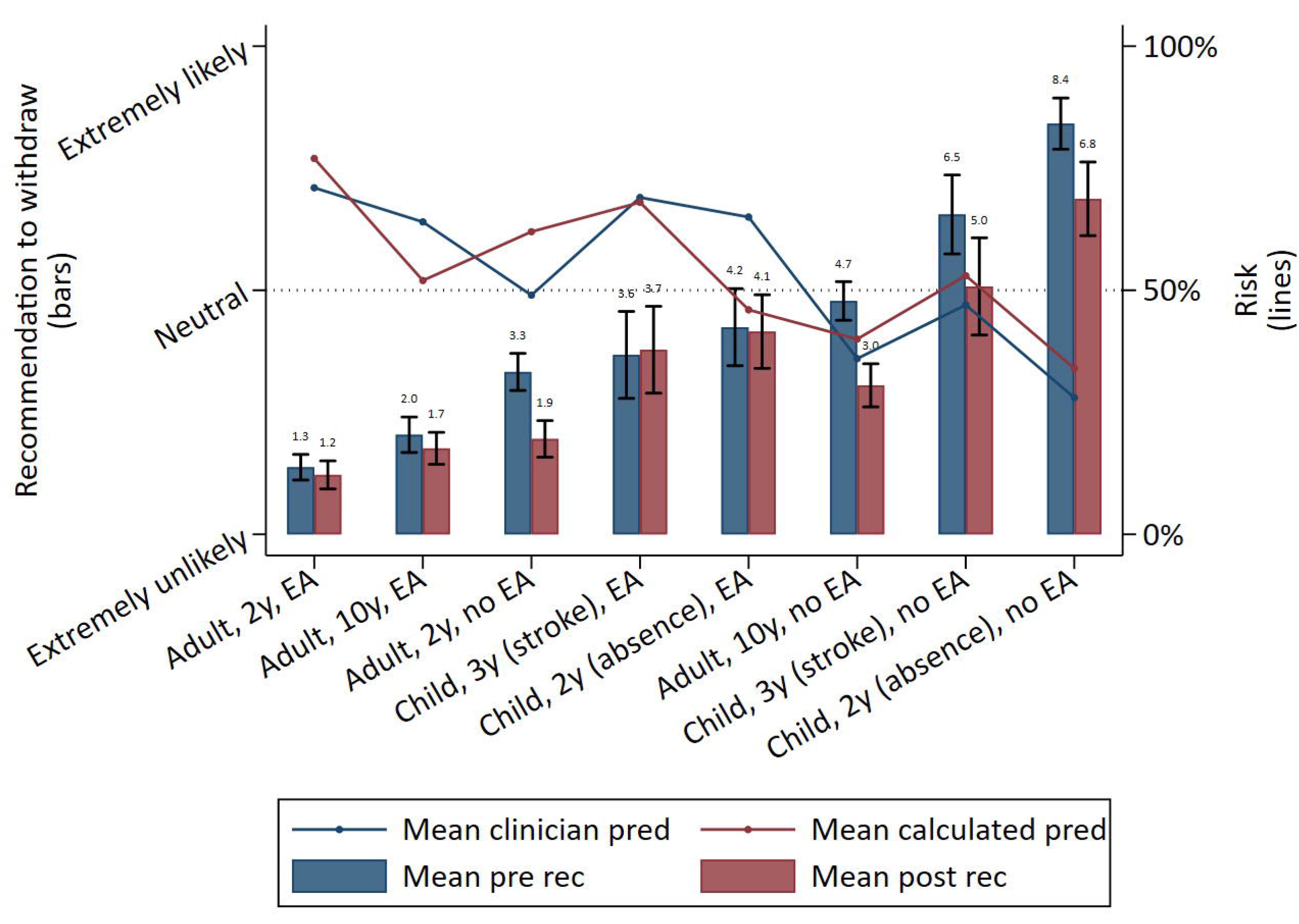
Recommendation to withdraw before (pre) versus after (post) viewing results of the calculator according to vignette, overlaid with post-withdrawal calculated predictions. Bars represent mean (+/- 95% CI) recommendations to withdraw before (blue) versus after (red) seeing calculated predictions and refer to the left y-axis. Lines represent mean estimations of two-year post-withdrawal clinical (blue) and calculated (red) risk predictions and refer to the right y-axis. *Interpretation*: Viewing calculated risks had no significant effect on recommendations in vignettes with epileptiform EEG abnormalities, but reduced recommendations for vignettes without epileptiform EEG abnormalities. Increasing recommendations to withdraw antiseizure medications corresponded with decreasing vignette risks. 2y: 2 years seizure-free; 3y: 3 years seizure-free: 10y: 10 years seizure-free; EA: epileptiform abnormalities; pred: prediction.

In general, recommendations to withdraw increased as the vignette’s risk decreased (Supplemental Figure 6). However, changes in recommendations (after versus before seeing the calculator) correlated only weakly with the degree to which clinicians over- versus under-predicted risk compared with calculated predictions (Spearman correlation coefficient 0.31, 95% CI 0.26-0.37; Supplemental Figure 7).

Numerous vignette characteristics influenced recommendations, with the strongest recommendation to withdraw for children without epileptiform abnormalities with side effects (8.6/10), and the strongest recommendation against withdrawing for adults with driving needs and no side effects (1.6/10) (Supplemental Figure 8). Epileptologists/clinical neurophysiologists were more likely to withdraw both before and after viewing calculated predictions compared with non-specialists for several vignettes, particularly for childhood cases (Supplemental Figure 9). After adjusting for all respondent and vignette characteristics, the strongest factor influencing recommendations was whether respondents treat only surgical patients (Figures e-10 and e-11), which predicted higher likelihood for recommending withdrawal (8.6/10, 95% CI 8.0/10-9.2/10; versus respondents who treat some surgical patients 3.2/10, 95% CI 3.0/10-3.5/10; p<0.05). Though, there were only 10 datapoints from surgical-only respondents, of the total 3,181 datapoints in this regression. Vignette characteristics increasing the likelihood of recommending withdrawal were pediatric or surgical vignettes, side effects, normal EEG, longer seizure-free period, not seeing the calculator, and absence of a lesion (p<0.05). For example, an epileptiform EEG reduced recommendations on average by 1.9 (95% CI 1.7 to 2.1) points on a 10-point scale.

### Frequency/barriers of using calculators

Of 279 respondents answering whether they use the Lamberink online risk calculator, responses were: 151 (54%) never, 66 (24%) less than half the time, 19 (7%) half the time, 19 (7%) more than half the time, and 24 (9%) always. Of the 128 respondents who ever use the calculator, respondents find it: 20 (16%) very useful, 29 (23%) useful, 41 (32%) somewhat useful, 21 (16%) a little useful, 1 (1%) not at all useful, and 16 (13%) “I don’t know.” Of the 280 responding about barriers to using the calculator, top barriers included being unsure regarding its accuracy (122; 44%), not integrated into the medical record (116; 41%), unsure if it applies to a given patient (112; 40%), unaware that such a calculator existed (103; 37%), unsure how to find it (89; 32%), not enough time to use it during visits (61; 22%), and 14 (5%) indicated there are no barriers. Free-text comments included the importance of patient preferences, that calculated risk may not influence their decisions unless calculations were very close to 0% or 100%, and difficulty for patients thinking in terms of chance rather than certainty.

## Discussion

We performed an international survey exploring how neurologists estimate seizure risk and make recommendations about ASM withdrawal in patients with well-controlled epilepsy. While mean clinician predictions were mostly close to calculated predictions, we found wide variation in clinician predictions and recommendations within all vignettes. Respondents overestimated risk in the lowest-risk vignettes with epileptiform abnormalities (i.e., childhood absence epilepsy; ten-year seizure-free adult; surgical patients) and overestimated the influence of EEG abnormalities compared with best-available calculation. Viewing calculator results reduced recommendations to withdraw for vignettes with no epileptiform abnormalities but made little difference in recommendations for vignettes with epileptiform abnormalities.

Variation in recommendations is not unexpected. Current science does not inform any single optimal risk cutoff, all clinicians may have different thresholds tailored to their population, and prior work has suggested that epileptologists might be more likely to withdraw ASMs perhaps related to greater comfort with such decisions.[29] However, wide variation in risk estimation when shown the same clinical data is concerning, given two clinicians with the same risk threshold seeing similar patients may reach very different conclusions. This encourages continuing to develop accurate seizure risk prediction tools to better standardize risk estimation. One may have hypothesized “more accurate” risk estimation in clinicians with greater subspecialization or years of experience, but this was not the case. Rather, our data suggested that risk estimation may not align with best-available risk calculators particularly for certain types of patients, rather than for certain provider types.

Our results may question certain common practices regarding ASM withdrawal. For example, two-thirds of respondents always obtain EEGs before withdrawal. However, whereas clinicians estimated that epileptiform EEG abnormalities increased adjusted risk by 30-40% and epileptiform EEG abnormalities had a modest influence on recommendations, calculated predictions suggested that epileptiform EEG abnormalities increased risk by only approximately 10-15%. Thus, always obtaining an EEG may not be required, depending on what absolute risk difference a clinician feels is clinically meaningful. Several free-text responses expressed the importance of EEGs yet also underscored that some clinicians would never advise withdrawal until seizure risk dropped very close to 0%, which is not achievable even with a normal EEG, even for patients who have never had a seizure.[30] Furthermore, our results suggest that clinicians may be too cautious regarding postsurgical ASM withdrawal. Calculated predictions for postoperative vignettes were about 20%-30% lower than clinician predictions, often about 50% lower than clinician predictions. Furthermore, calculated predictions suggested that awaiting two years compared with six months of seizure-freedom reduced the patient’s risk only by about 3%. Thus, for a postoperative patient who plans to eventually withdraw ASMs, early withdrawal may be justified, despite average recommendations being in the range of “extremely unlikely” to “unlikely” for our surgical cases. The TimeToStop study in children showed that ASM withdrawal as soon as 6 months after epilepsy surgery does not influence long-term seizure outcome,[31] which could likewise apply to adults.

Viewing calculated risk particularly reduced recommendations without epileptiform EEG abnormalities. For example, the mean clinician predicted post-withdrawal risk was nearly identical to the Lamberink calculated risk for a ten-year seizure-free adult. Nonetheless, viewing calculated risk still reduced recommendations to withdraw. Perhaps making decisions according to clinician’s intuitive understanding that this is a relatively low-risk case may lead to more liberal recommendations, whereas requiring clinicians to articulate probabilities may lead to more conservative recommendations.

Almost half of respondents ever use the available online risk calculator, and 40% reported being unsure whether it is accurate. We recognize that no there is no single “gold-standard” model, which is why we took caution to present numerous methods to calculate risk for each case. Developing a model with high accuracy and widespread applicability represents a key, challenging, research goal. Even if a calculator predicted risk perfectly, other implementation barriers still include dissemination and integration into clinicians’ workflow, deciding whether and how to communicate complex probabilities to patients who may have low numeracy,[32] and most fundamentally, understanding what probability threshold, if any, merits ASM withdrawal.

Our work has limitations. The response rate was low. While our international survey reached a diverse audience, high response rates are difficult to achieve during physician research.[33] Still, our inverse probability of selection weighting accounted for likely the most pertinent clinician variables influencing study participation (e.g., epilepsy specialization, practice setting, seniority/experience, etc.) without changing conclusions. We also did not survey non-neurologists. Second, we did not allow the option of reducing doses or switching ASMs when we presented vignettes. Our questions mimicked existing guidelines on this topic which present withdrawal as a dichotomous decision. Adding considerations of reducing or switching ASMs into our survey would have added further considerable complexity and length. And above, only so many vignettes are feasible within any survey.

## Conclusions

Respondents provided highly variable clinical risk predictions and recommendations for ASM withdrawal. Wide variation encourages efforts at developing more evidence-based approaches to determining which patients benefit from continued ASM treatment versus withdrawal and future efforts developing point-of-care seizure risk prediction tools integrated into the electronic medical record. Clinicians overestimated the influence of epileptiform abnormalities, seizure-free duration, and withdrawal risk for surgical patients compared with calculated results, which may question widespread ordering of EEGs or time-based seizure-free thresholds and may encourage using calculated predictions in such scenarios. Viewing calculator results reduced recommendations to withdraw particularly for cases with a normal EEG. Future research is needed regarding how low seizure risk should be before ASM withdrawal.

## Supporting information

CROSS checklist

Survey instrument

## Data Availability

The American Academy of Neurology is the sole owner of the raw data. Data can only be shared with a data use agreement. All data produced in the present work are contained in the manuscript.

## Ethical publication statement

We confirm that we have read the Journal’s position on issues involved in ethical publication and affirm that this report is consistent with those guidelines.

## Author contributions

S.W.T. conceived of and designed the study, executed the statistical analysis, and wrote the manuscript. C.E. Rheaume collected the data. All authors assisted with designing the survey, interpretation, and manuscript editing.

## Acknowledgments

We are deeply grateful to each neurologist who took the time to contribute data to this study.

---

**Supplemental Table 1:** Vignettes.

| **Vignette** | **Adult** | **Adult** | **Surgical, MTS** | **Surgical, MTS** | **Surgical, FCD** | **Surgical, FCD** | **Child, absence** | **Child, stroke** |
| --- | --- | --- | --- | --- | --- | --- | --- | --- |
| **Sz-free** | 2y | 10y | 6m | 2y | 6m | 2y | 2y | 3y |
| **Age, onset** | 26 | 26 | 15 | 15 | 15 | 15 | 8 | 5 |
| **Age, last sz** | 28 | 28 | 30 | 30 | 30 | 30 | 9 | 8 |
| **Age, current** | 30 | 38 | 30.5 | 32 | 30.5 | 32 | 11 | 11 |
| **Sex** | M | M | M | M | F | F | F | F |
| **Duration, sz's, y** | 2 | 2 | 15 | 15 | 15 | 15 | 1 | 3 |
| **Duration, sz-free, y** | 2 | 10 | 0.5 | 2 | 0.5 | 2 | 2 | 3 |
| **# seizures** | 6 | 6 | Innum | Innum | Innum | Innum | Innum | 6 |
| **Semiology** | FBTC | FBTC | FBTC | FBTC | FBTC | FBTC | Abs.+GTCs | FM |
| **Etiology** | UK | UK | MTS | MTS | FCD | FCD | UK | Stroke |
| **Surgical** | N | N | Y | Y | Y | Y | N | N |
| **FH** | N | N | N | N | N | N | N | N |
| **FS** | Y | Y | Y | Y | N | N | N | N |
| **DD** | N | N | N | N | Y | Y | N | Y |
| **ASM** | Lev | Lev | Lev | Lev | Phen | Phen | VPA | Lev |
| **# responded** | 136 | 130 | 73 | 73 | 76 | 76 | 40 | 38 |

Sz-free: duration since last seizure; 2y: 2 years seizure-free; 10y: 10 years seizure-free; 6m: 6 months seizure-free; sz: seizure; Innum: innumerable; FBTC: focal to bilateral tonic-clonic; Abs: absence; GTCs: generalized tonic-clonic seizures; FM: focal motor; MTS: mesial temporal sclerosis; FCD: focal cortical dysplasia; UK: unknown; FH: family history; FS: febrile seizures; DD: developmental delay; ASM: antiseizure medication; Lev: levetiracetam; Phen: phenytoin; VPA: valproic acid; # responded: number of neurologists who provided at least one clinician prediction or recommendation for each vignette

**Supplemental Figure 1.**
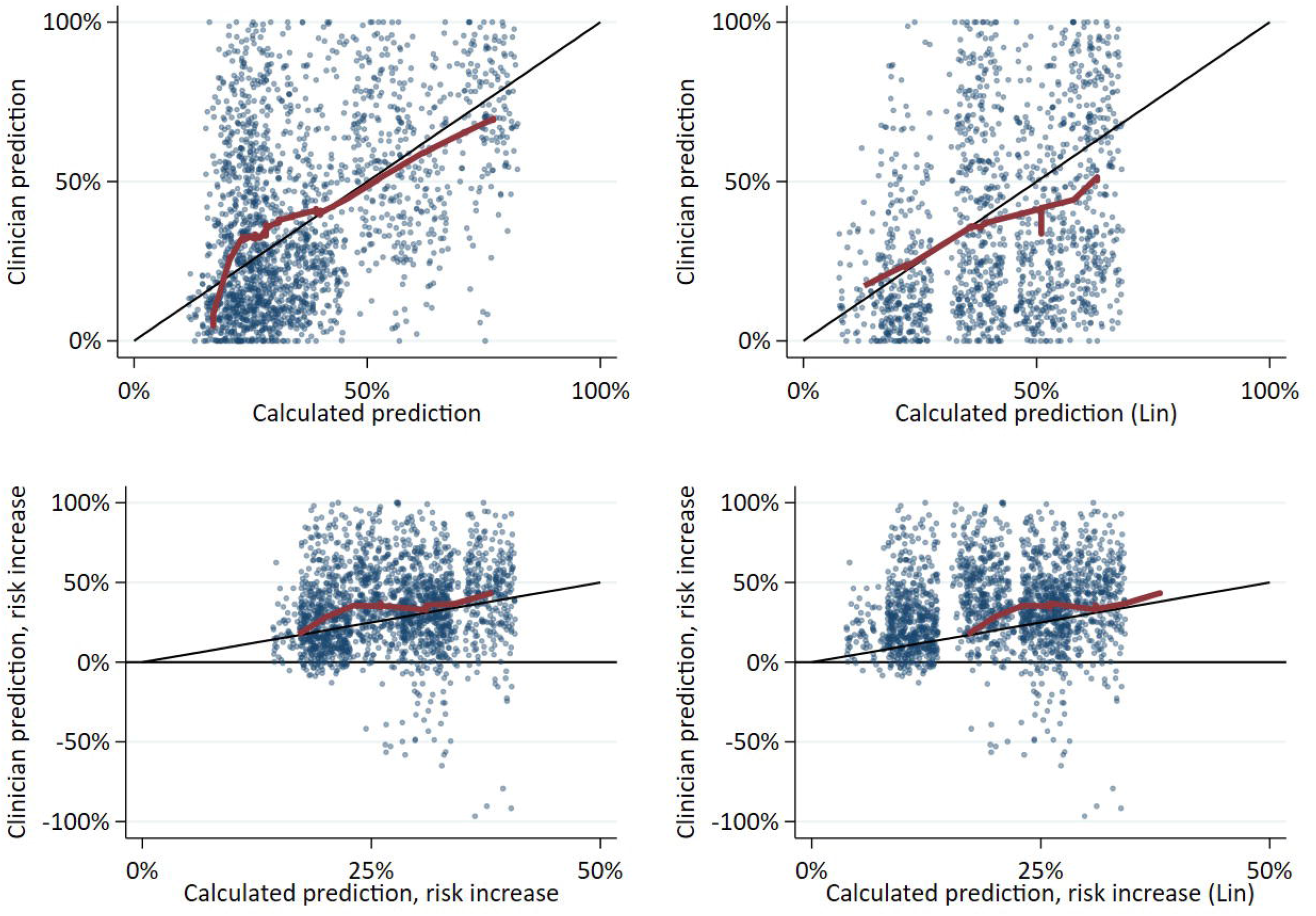
Clinical (y-axis) versus calculated (x-axis) two-year seizure risk predictions (top) and absolute risk increase due to withdrawal (bottom). Left: Risks on the x-axis were calculated using the Lamberink model for medical vignettes and the Ferreira-Atuesta model for surgical vignettes. Right: Risks on the x-axis were calculated using the Lin model for medical vignettes only. All points have a small amount of jitter added for visualization. Black 45° lines would represent perfect correlation. Red loess curves nonparametrically fit the data. Note in the bottom plots that the 5% of dots below the y-axis may have represented respondents who did not understand the instructions (all dots should in theory be above 0 given withdrawal that increases risk). *Interpretation:* There was wide variation in clinical versus calculated predictions and treatment effects.

**Supplemental Figure 2.**
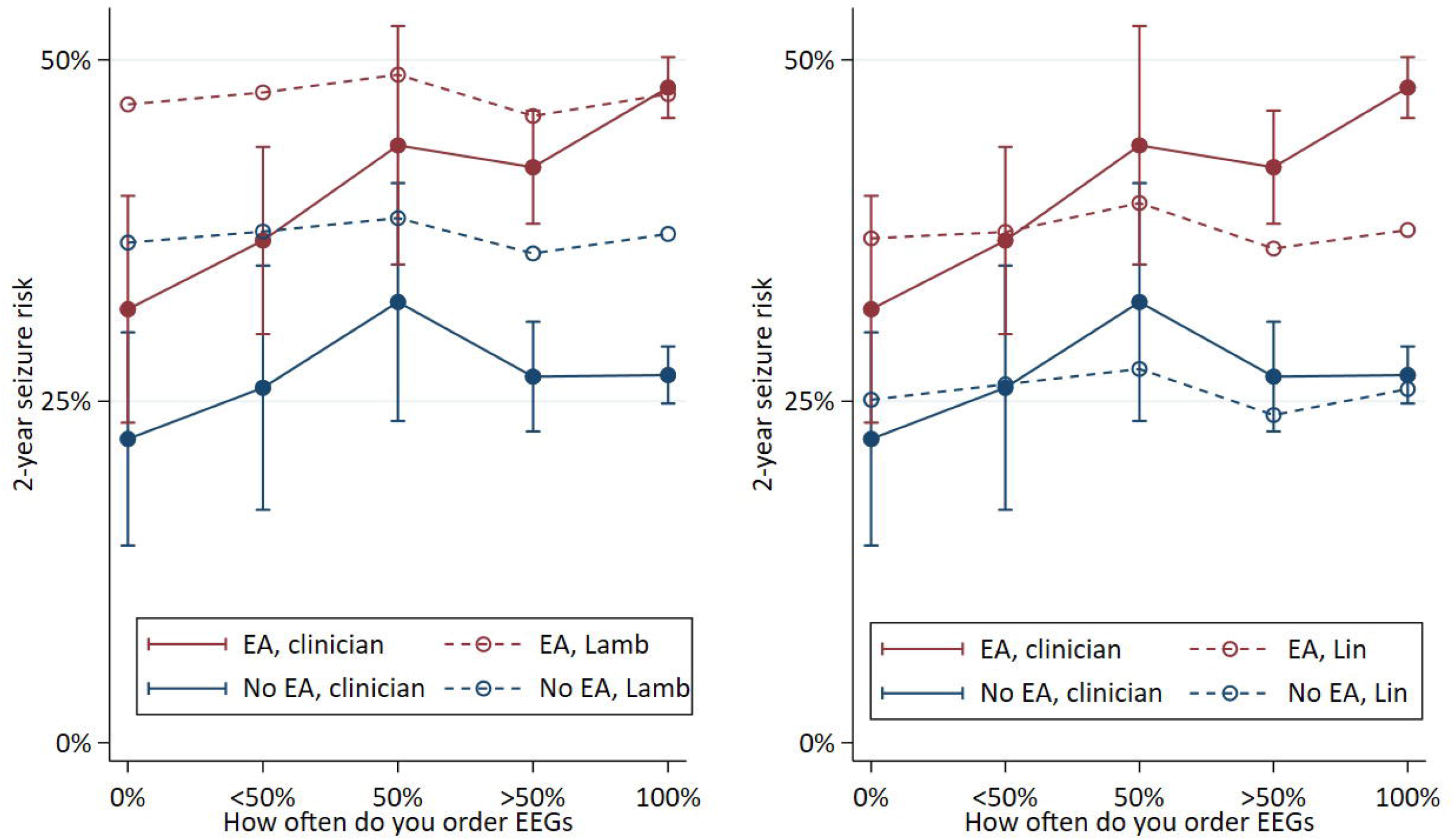
Influence of epileptiform EEG abnormalities (EA) on post-withdrawal clinician and calculated predictions. We compared how much clinicians (solid red versus blue lines) versus calculators (dashed red versus blue lines) predicted that seizure risk is increased due to EAs, averaged over medical vignettes. Note the solid lines in the left and right panels are identical. What differs between the panels is that we displayed Lamberink calculations in the left panel and Lin calculations in the right panel, given we acknowledge that there is no single one gold standard calculation method and thus wished to show data both ways. We also went one step further – because we hypothesized that clinicians who most often order EEGs might believe EEGs increase risk more than clinicians who order EEGs less often, we stratified by how often respondents indicated they order EEGs when facing withdrawal decisions. Error bars represent 95% confidence intervals. *Interpretation*: On average, respondents overestimated the influence of epileptiform abnormalities, particularly among those who always order EEGs when considering withdrawal.

**Supplemental Figure 3.**
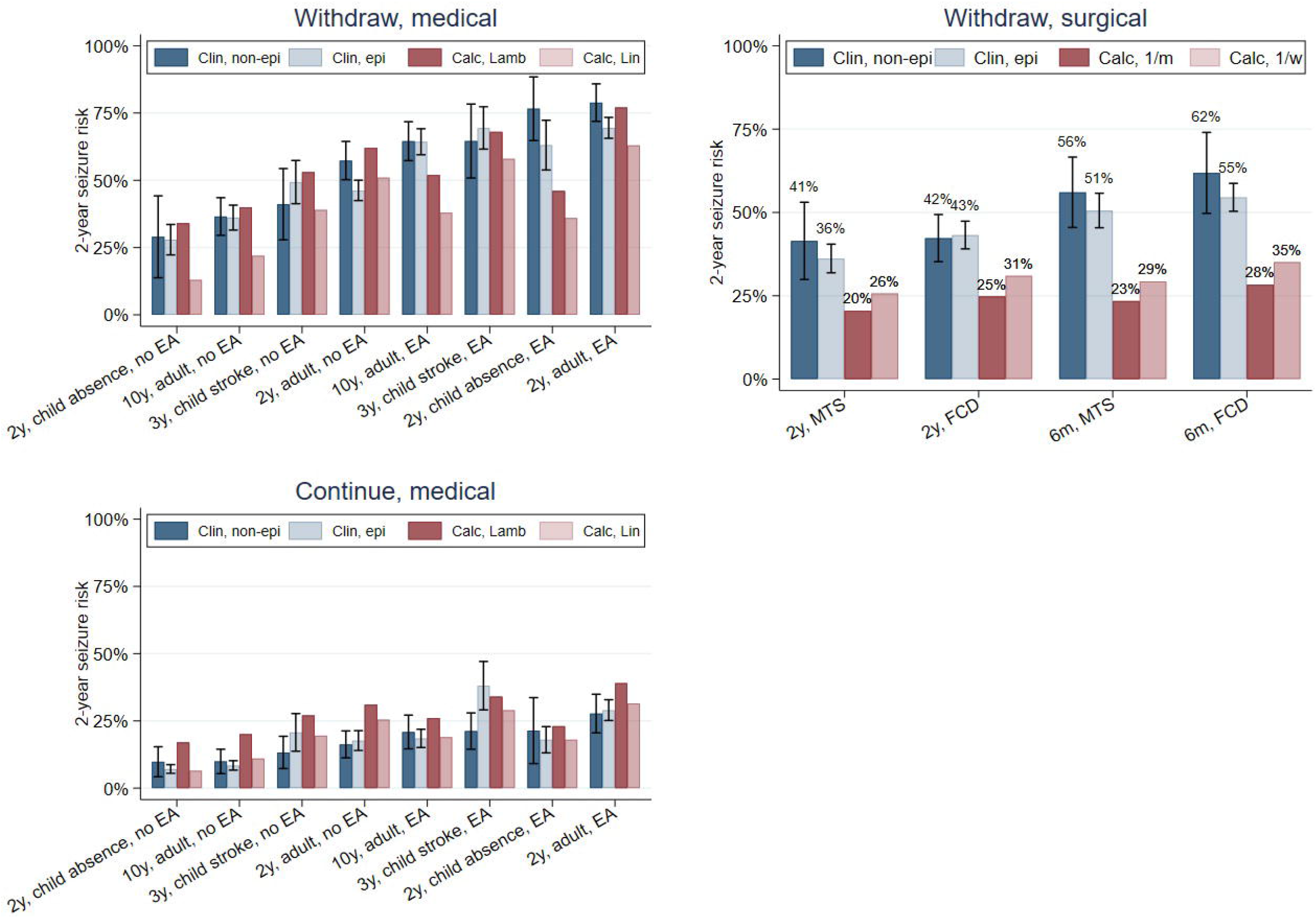
Mean and 95% confidence intervals for clinician versus calculated predictions of two-year seizure relapse risk by vignette, stratified by respondent specialization. This is displaying similar information as Figure 2, except we have stratified clinician predictions according to whether respondents specialize in epilepsy or clinical neurophysiology (collectively referred to as “epi” in the legend). *Interpretation:* Mean clinician predictions were mostly similar between specialists versus non-specialists.

**Supplemental Figure 4.**
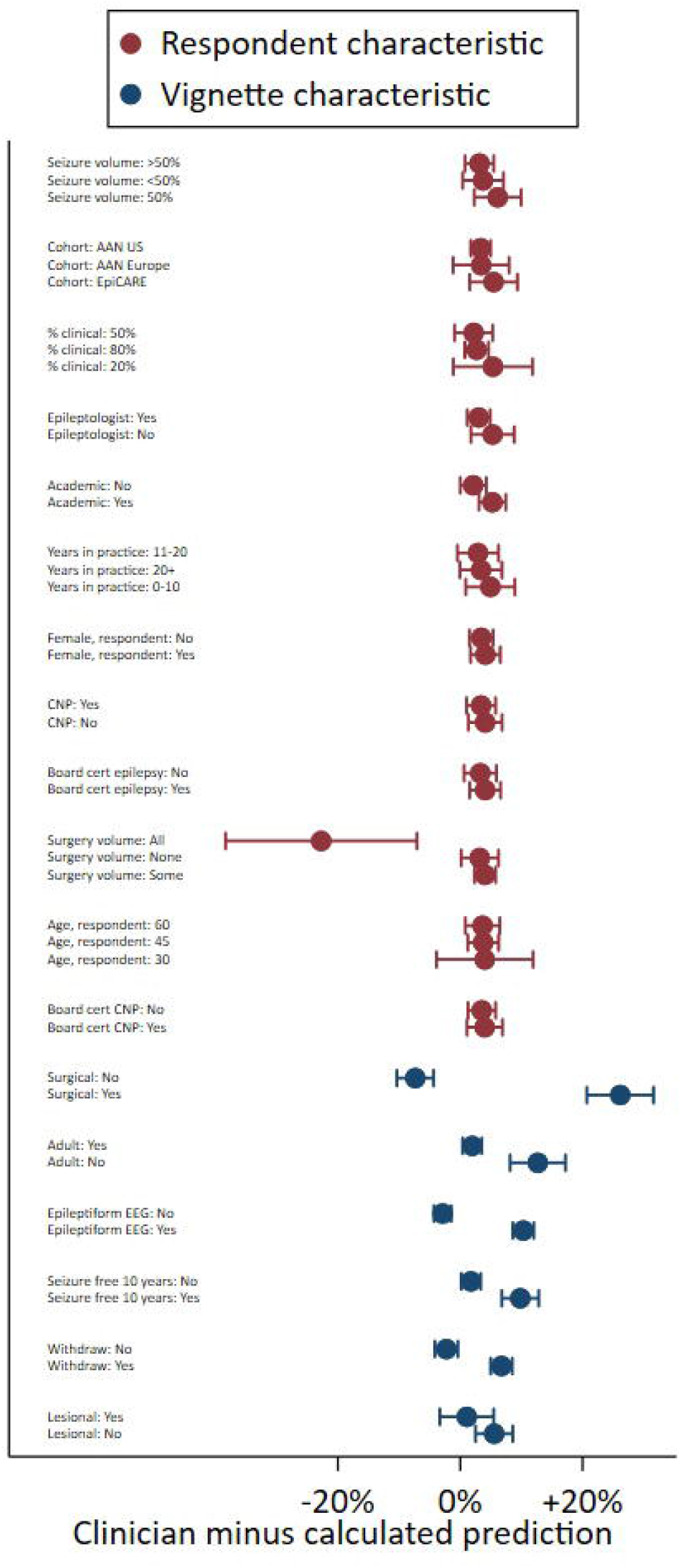
Adjusted mean and 95% confidence intervals for clinical minus calculated (Lamberink) predictions according to respondent and vignette characteristics. These data were produced from a single model that pooled all responses. *Interpretation:* Most respondent characteristics influenced differences very little. Differences were more largely driven by vignette characteristics.

**Supplemental Figure 5.**
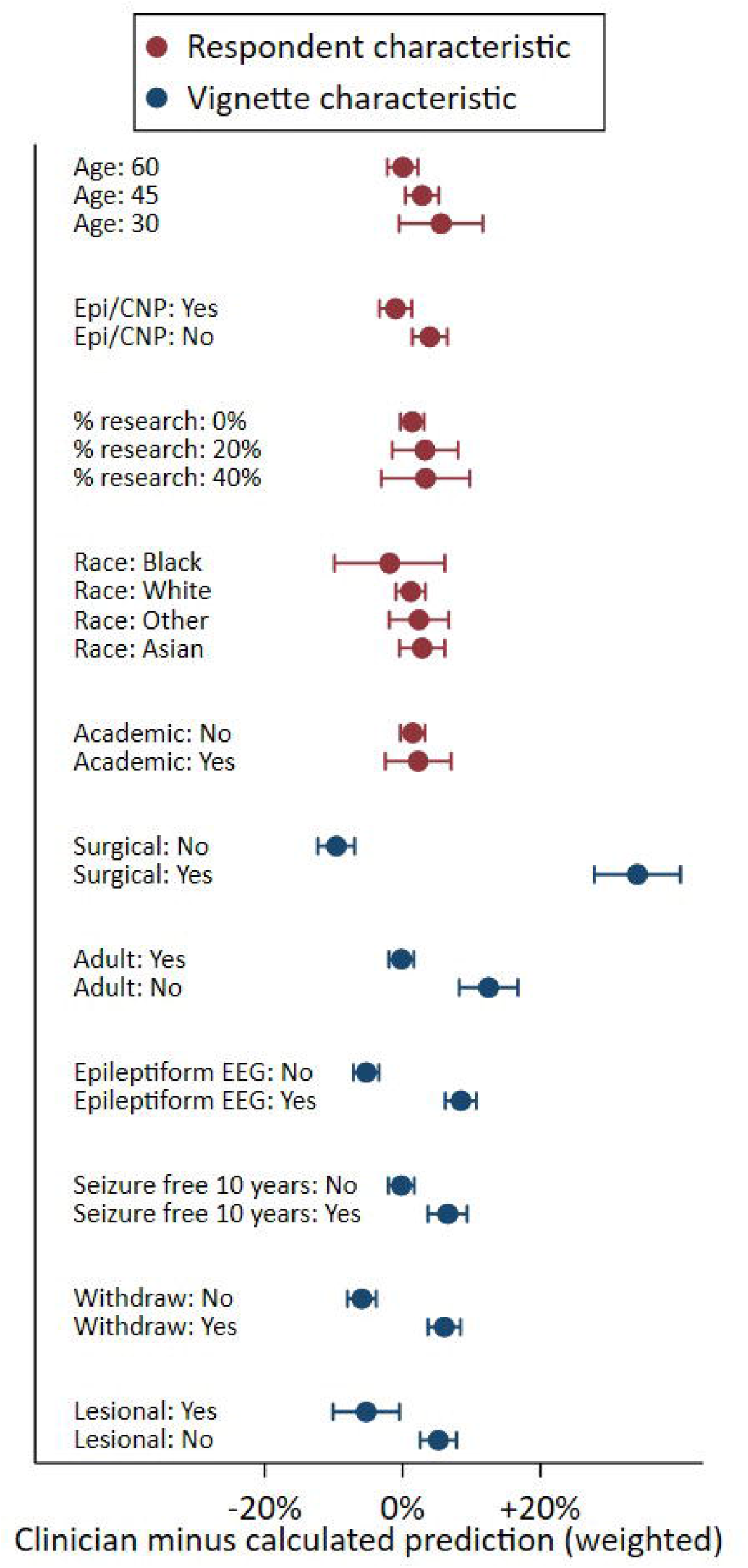
Adjusted clinician minus calculated (Lamberink) risks according to respondent and vignette characteristics: weighted. Mean +/- 95% confidence interval. *Interpretation*: Similar to Figure 3.

**Supplemental Figure 6.**
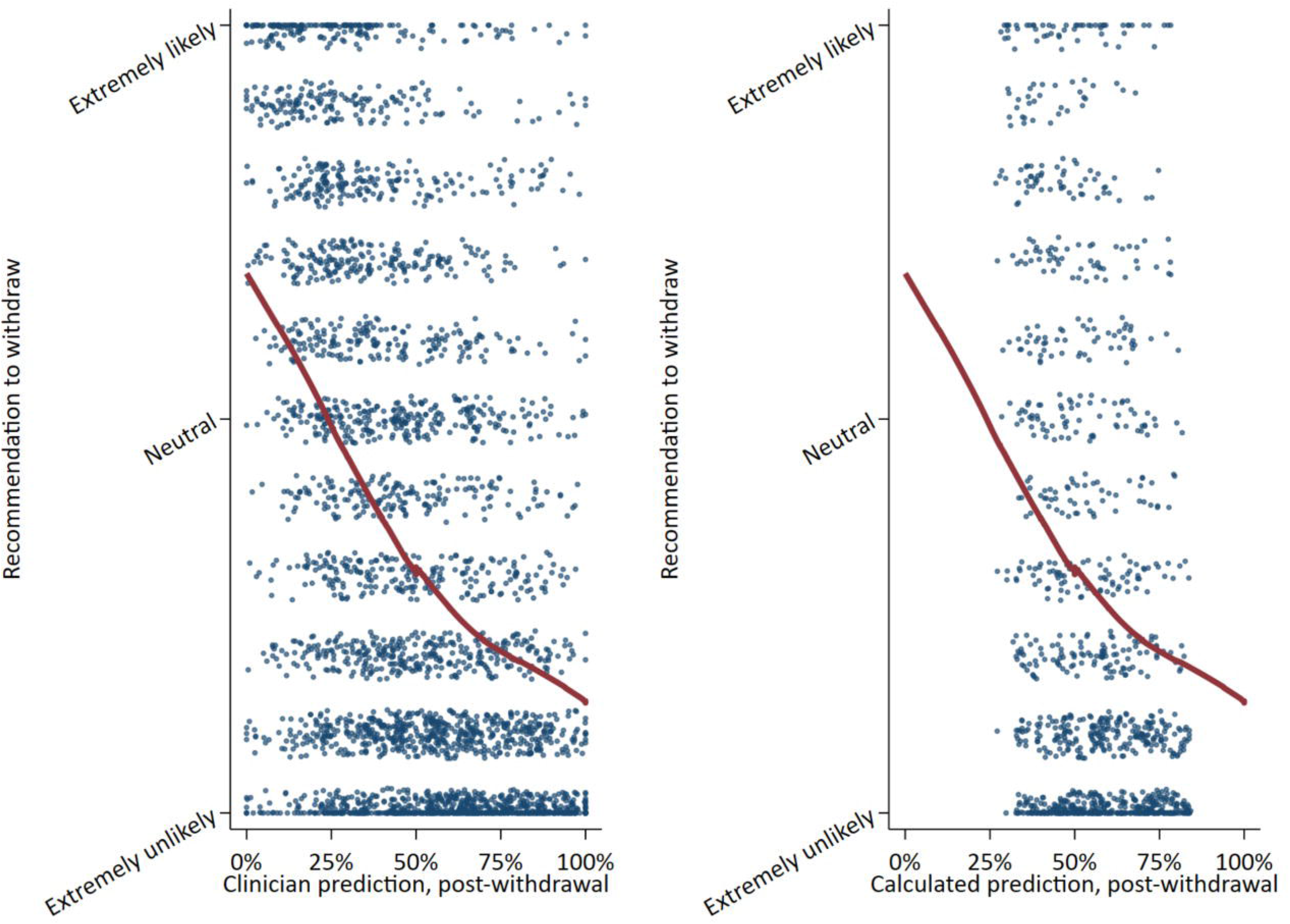
Recommendation to withdraw according to clinician (left) and calculated (right) predictions. Red lines represent lowess curves fitting the data. *Interpretation:* Recommendations to withdraw increased as risk decreased. The range of clinician predictions was wider than the range of calculated predictions, and the recommendations ranged the entire spectrum from extremely unlikely to extremely likely throughout the risk continuum.

**Supplemental Figure 7.**
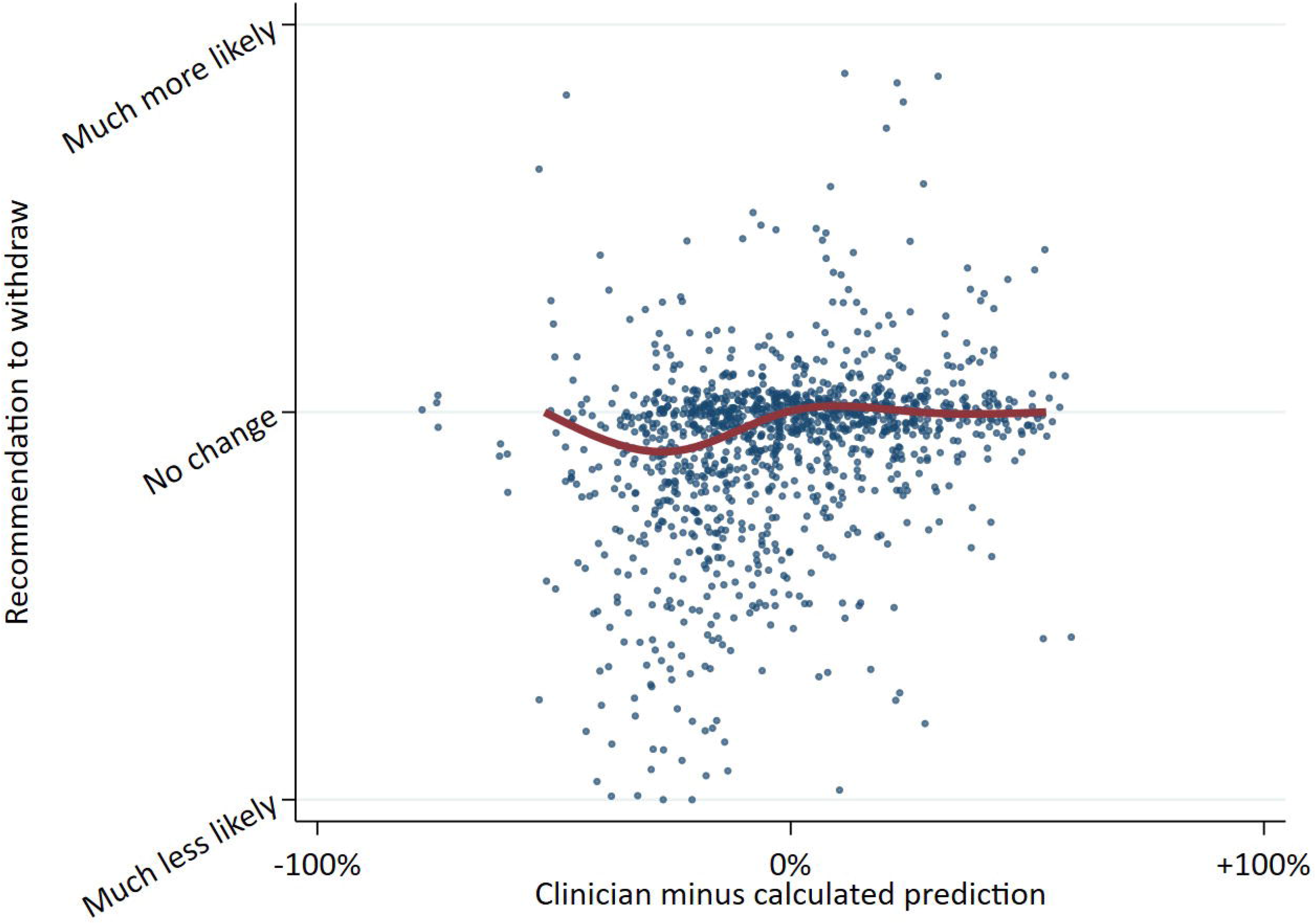
Change in recommendation according to clinician minus calculated prediction. The y-axis represents how much respondents’ recommendations changed after seeing calculated risks (higher = more likely after seeing calculated result). The x-axis represents how much higher or lower clinician predictions were compared with calculated predictions (higher = clinicians predicted a higher 2-year post-withdrawal seizure risk compared with calculated predictions). *Interpretation:* One might have hypothesized there would have been a positive slope if respondents guessing risks higher than the calculator would then have revised their recommendations to more strongly recommend withdrawal if the calculator influenced recommendations. However, only weak correlation (Spearman’s rho 0.31, 95% CI 0.26-0.36) may mean that respondents did not factor calculator results consistently into their recommendations.

**Supplemental Figure 8.**
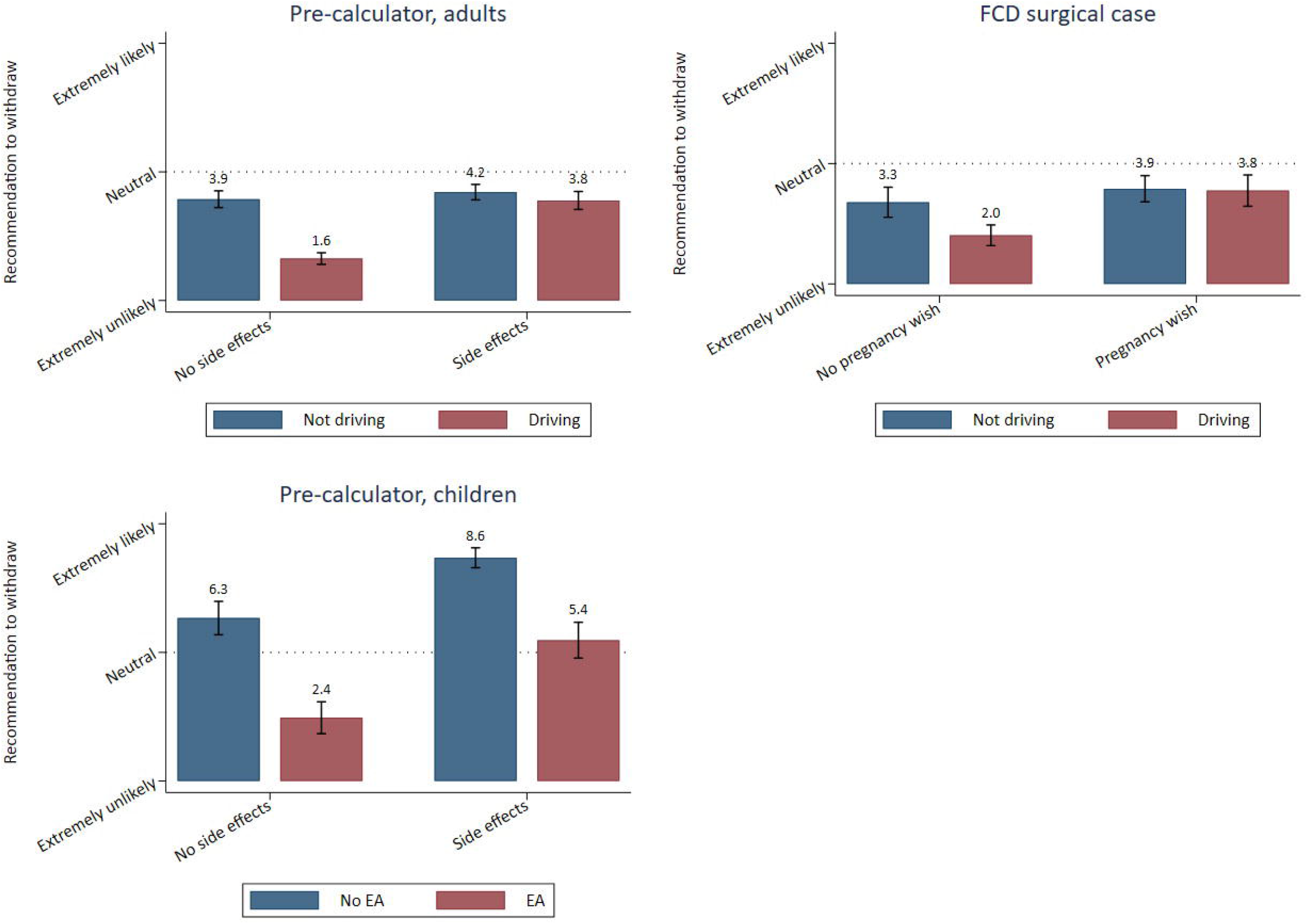
Recommendations (mean and 95% confidence interval) to withdraw before seeing calculated predictions, according to additional vignette factors. *Interpretation:* Driving needs decreased recommendations to withdraw for patients without side effects but had little influence on recommendations for patients with side effects. Similarly, driving needs decreased recommendations to withdraw for patients without pregnancy wish but had little influence on recommendations for patients with a pregnancy wish. In children, side effects increased recommendations to withdraw as well. FCD: focal cortical dysplasia; EA: epileptiform abnormalities.

**Supplemental Figure 9.**
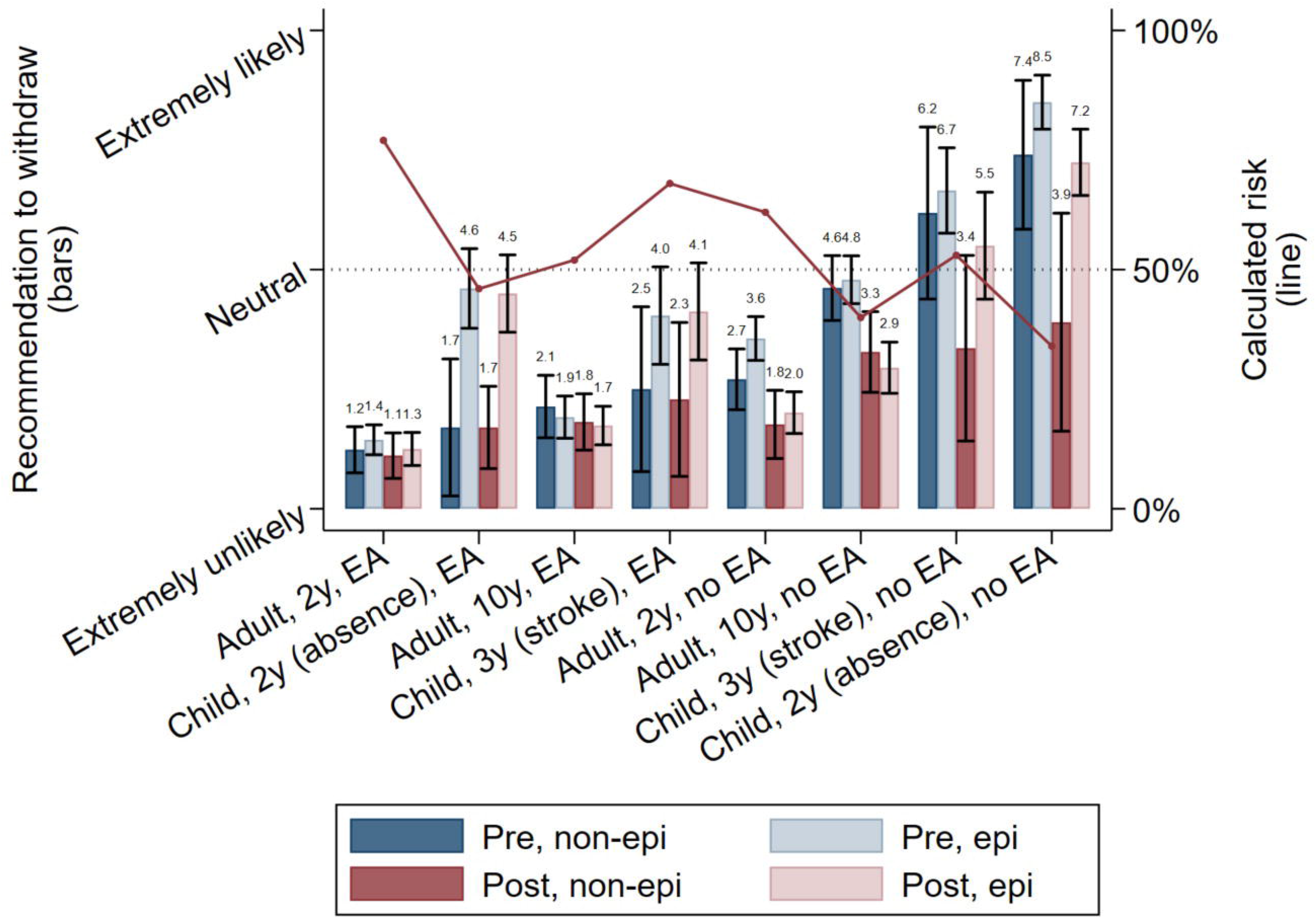
Recommendation to withdraw before (pre) versus after (post) viewing results of the calculator according to vignette, stratified by subspecialization. This is displaying similar information as Figure 5 except we have stratified clinician predictions according to whether respondents specialize in epilepsy or clinical neurophysiology (collectively referred to as “epi” in the legend). *Interpretation:* Specialists were more likely than non-specialists to recommend medication withdrawal for several vignettes, particularly for several childhood cases.

**Supplemental Figure 10.**
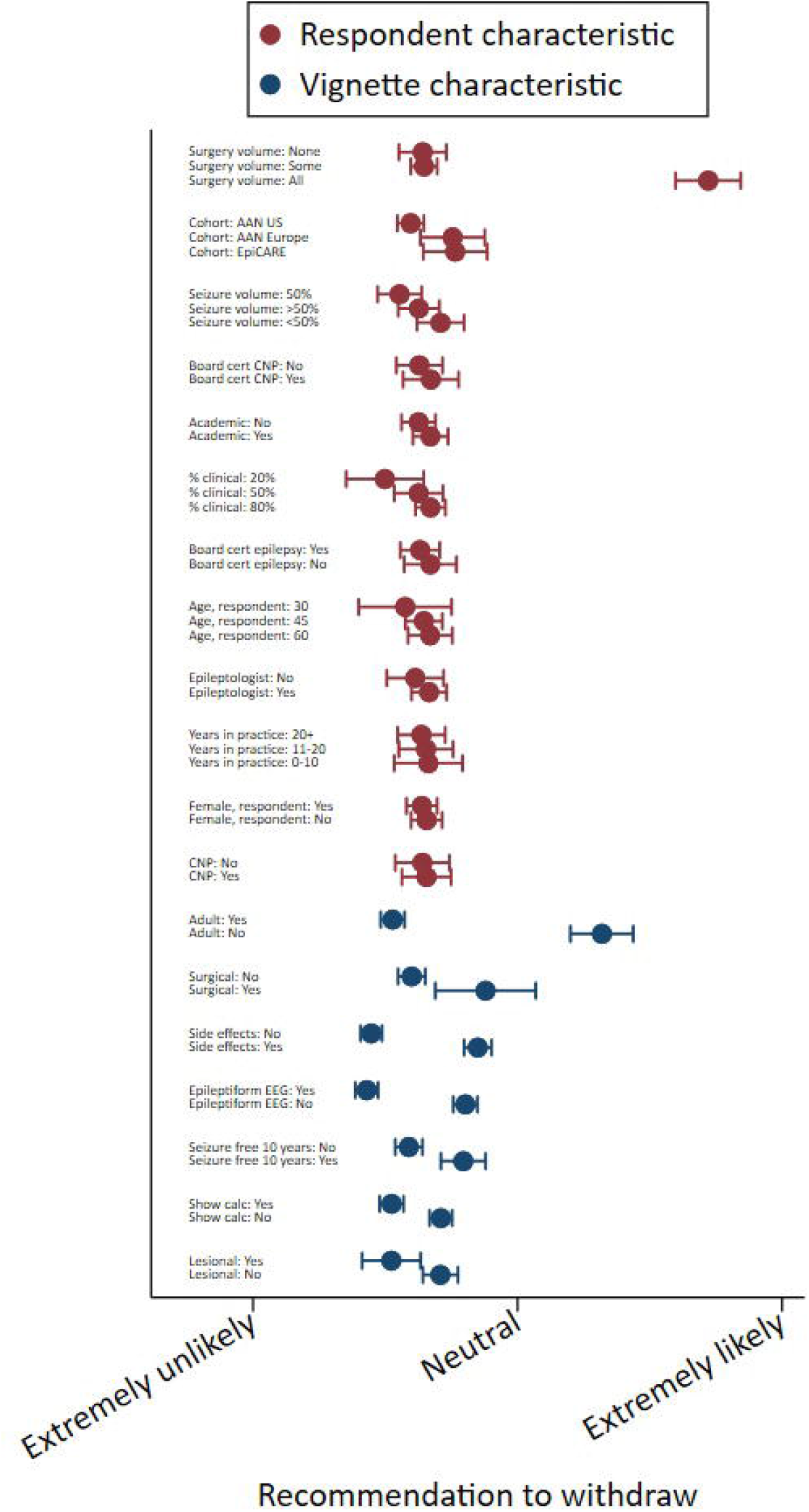
Adjusted recommendations according to respondent and vignette characteristics. Mean +/- 95% confidence interval. *Interpretation:* Respondents were much more likely to recommend withdrawal if they treat only surgical patients, but other respondent characteristics made little difference. Vignette characteristics increasing the likelihood of recommending withdrawal were pediatric or surgical vignettes, side effects, normal EEG, longer seizure-free period, not seeing the calculator, and absence of a lesion. AAN: American Academy of Neurology; CNP: clinical neurophysiology.

**Supplemental Figure 11.**
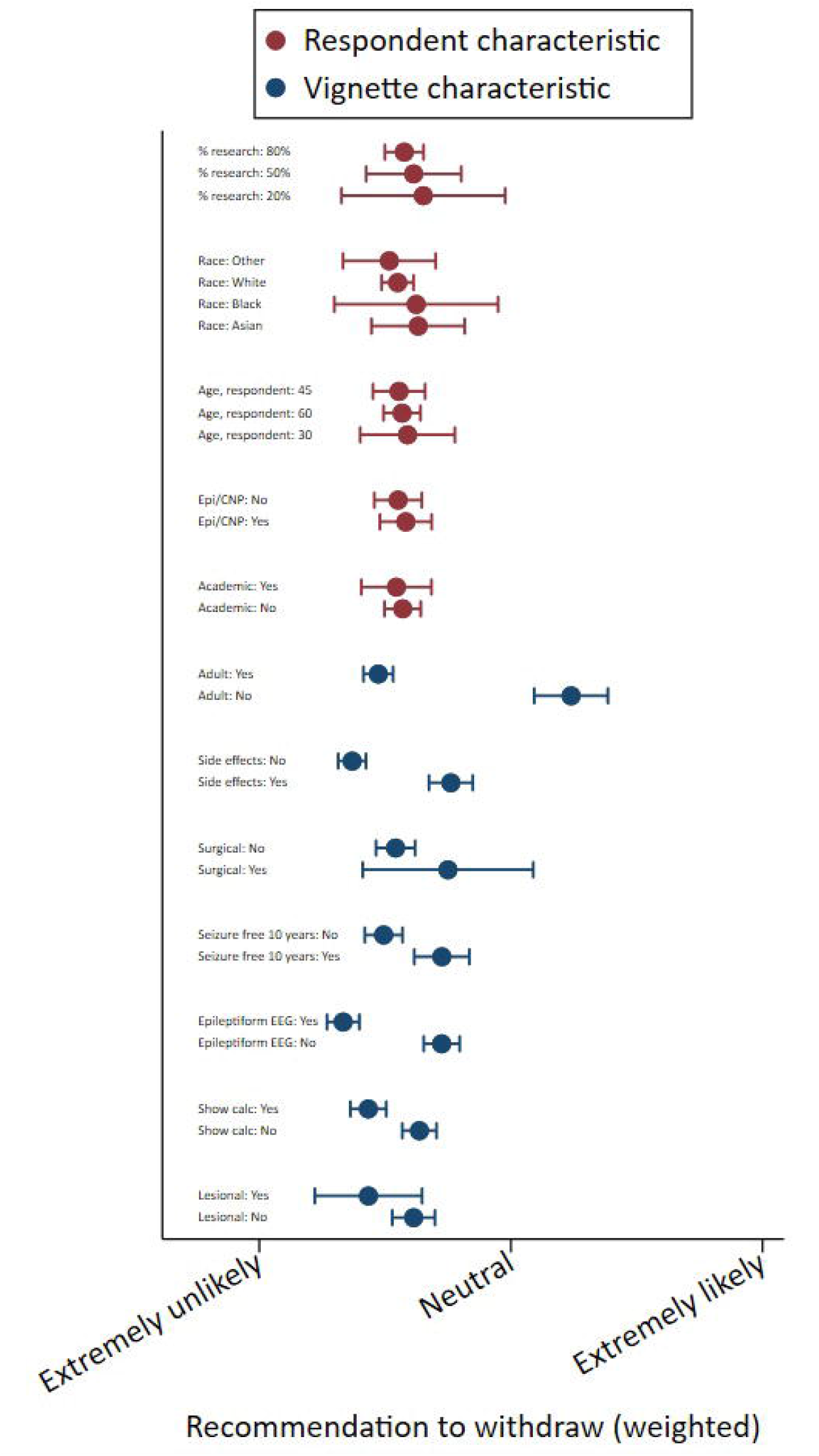
Adjusted recommendations according to respondent and vignette characteristics (weighted). *Interpretation*: Similar to Figure 6.

