## Supplementary material for "Antiseizure medication withdrawal risk estimation and recommendations: a survey of American Academy of Neurology and EpiCARE members": Survey instrument

Block: A: Screening Questions (8 Questions)

Branch: New Branch

If

If Are you a neurologist? No Is Selected

Or In the past year, which describes your practice setting? Mark all that apply. I do not have a clinical practice Is Selected

Or In the past year, how many of your patients were seen for seizure evaluation/management? None Is Selected

EndSurvey: Advanced

Branch: New Branch

If

If In the past year, which best describes your epilepsy/seizure patients? Mostly adult (18 years of age or older) patients Is Selected

And In the past year, how many of the patients that you are managing for epilepsy had undergone epile... Some Is Selected

BlockRandomizer: 1 - Evenly Present Elements

Group: Patient B1- Ad Med

Standard: B1A Ad Med Pt 1 Vignette A: Risk Est, M, 26 28 30, F&BTC, wo EA (5 Questions)

Standard: B1B Ad Med Pt 1 Vignette B: Risk Est, M, 26 28 30, F&BTC, w EA (4 Questions)

Branch: New Branch

If

If Here is the first patient.Sex: maleFirst seizure: 26 years old Last seizure: 28 years old (Durati... Is Displayed

Standard: B1C Ad Med Pt 1 Vignette C: Risk Given, M, 26 28 30, F&BTC, wo EA (3 Questions)

Standard: B1D Ad Med Pt 1 Vignette D: Risk Given: M, 26 28 30, F&BTC, w EA (3 Questions)

Group: Patient B2- Ad Med

Standard: B2A Ad Med Pt 2 Vignette A: Risk Est, M, 26 28 38, F&BTC, wo EA (5 Questions)

Standard: B2B Ad Med Pt 2 Vignette B: Risk Est, M, 26 28 38, F&BTC, w EA (4 Questions)

Branch: New Branch

If

If Here is the first patient.  Sex: maleFirst seizure: 26 years oldLast seizure: 28 years old (Durat... Is Displayed

Standard: B2C Ad Med Pt 2 Vignette C: Risk Given, M, 26 28 38, F&BTC, wo EA (3 Questions)

Standard: B2D Ad Med Pt 2 Vignette D Risk Given: M, 26 28 38, F&BTC, w EA (3 Questions)

BlockRandomizer: 1 - Evenly Present Elements

Group: Patient C1- Ad Surg

Standard: C1A Ad Surg Pt 1 Vignette A: M, 30, F&BTC, 6M, wo EA (6 Questions)

Standard: C1B Ad Surg Pt 1 Vignette B: M, 30, F&BTC, 6M, w EA (4 Questions)

Standard: C1C Ad Surg Pt 1 Vignette C: M, 30, F&BTC, 2Y, wo EA (4 Questions)

Standard: C1D Ad Surg Pt 1 Vignette D: M, 30, F&BTC, 2Y, w EA (4 Questions)

Group: Patient C2

Standard: C2A Ad Surgery Pt 2 Vignette A: F, 30, F&BTC, 6M, wo EA (6 Questions)

Standard: C2B Ad Surgery Pt 2 Vignette B: F, 30, F&BTC, 6M, w EA (4 Questions)

Standard: C2C Ad Surgery Pt 2 Vignette C: F, 30, F&BTC, 2Y, wo EA (4 Questions)

Standard: C2D Ad Surgery Pt 2 Vignette D: F, 30, F&BTC, 2Y, w EA (4 Questions)

Branch: New Branch

If

If In the past year, which best describes your epilepsy/seizure patients? Mostly adult (18 years of age or older) patients Is Selected

And In the past year, how many of the patients that you are managing for epilepsy had undergone epile... All Is Selected

BlockRandomizer: 1 - Evenly Present Elements

Group: Patient C1- Ad Surg

Standard: C1A Ad Surg Pt 1 Vignette A: M, 30, F&BTC, 6M, wo EA (6 Questions)

Standard: C1B Ad Surg Pt 1 Vignette B: M, 30, F&BTC, 6M, w EA (4 Questions)

Standard: C1C Ad Surg Pt 1 Vignette C: M, 30, F&BTC, 2Y, wo EA (4 Questions)

Standard: C1D Ad Surg Pt 1 Vignette D: M, 30, F&BTC, 2Y, w EA (4 Questions)

Group: Patient C2- Ad Surg

Standard: C2A Ad Surgery Pt 2 Vignette A: F, 30, F&BTC, 6M, wo EA (6 Questions)

Standard: C2B Ad Surgery Pt 2 Vignette B: F, 30, F&BTC, 6M, w EA (4 Questions)

Standard: C2C Ad Surgery Pt 2 Vignette C: F, 30, F&BTC, 2Y, wo EA (4 Questions)

Standard: C2D Ad Surgery Pt 2 Vignette D: F, 30, F&BTC, 2Y, w EA (4 Questions)

Branch: New Branch

If

If In the past year, which best describes your epilepsy/seizure patients? Mostly adult (18 years of age or older) patients Is Selected

And In the past year, how many of the patients that you are managing for epilepsy had undergone epile... None Is Selected

BlockRandomizer: 1 - Evenly Present Elements

Group: Patient B1- Ad Med

Standard: B1A Ad Med Pt 1 Vignette A: Risk Est, M, 26 28 30, F&BTC, wo EA (5 Questions)

Standard: B1B Ad Med Pt 1 Vignette B: Risk Est, M, 26 28 30, F&BTC, w EA (4 Questions)

Branch: New Branch

If

If Here is the first patient.Sex: maleFirst seizure: 26 years old Last seizure: 28 years old (Durati... Is Displayed

Standard: B1C Ad Med Pt 1 Vignette C: Risk Given, M, 26 28 30, F&BTC, wo EA (3 Questions)

Standard: B1D Ad Med Pt 1 Vignette D: Risk Given: M, 26 28 30, F&BTC, w EA (3 Questions)

Group: Patient B2 - Ad Med

Standard: B2A Ad Med Pt 2 Vignette A: Risk Est, M, 26 28 38, F&BTC, wo EA (5 Questions)

Standard: B2B Ad Med Pt 2 Vignette B: Risk Est, M, 26 28 38, F&BTC, w EA (4 Questions)

Branch: New Branch

If

If Here is the first patient.  Sex: maleFirst seizure: 26 years oldLast seizure: 28 years old (Durat... Is Displayed

Standard: B2C Ad Med Pt 2 Vignette C: Risk Given, M, 26 28 38, F&BTC, wo EA (3 Questions)

Standard: B2D Ad Med Pt 2 Vignette D Risk Given: M, 26 28 38, F&BTC, w EA (3 Questions)

Branch: New Branch

If

If In the past year, which best describes your epilepsy/seizure patients? Mostly adult (18 years of age or older) patients Is Selected

And In the past year, how many of the patients that you are managing for epilepsy had undergone epile... All Is Not Selected

Branch: New Branch

If

If In the past year, which best describes your epilepsy/seizure patients? Mostly pediatric (17 years of age or younger) patients Is Selected

And In the past year, how many of the patients that you are managing for epilepsy had undergone epile... All Is Not Selected

BlockRandomizer: 1 - Evenly Present Elements

Group: Patient D1 - Ped Med

Standard: D1A Ped Med Pt 1 Vignette A Risk Est: F, 8 9 11, A&GTC, wo EA (4 Questions)

Standard: D1B Ped Med Pt 1 Vignette B Risk Est: F, 8 9 11, A&GTC, w EA (3 Questions)

Branch: New Branch

If

If Here is the first patient. Sex: femaleFirst seizure: 8Last seizure: 9 (Duration of seizures: 1 ye... Is Displayed

Standard: D1C Ped Med Pt 1 Vignette C Risk Given: F, 8 9 11, A&GTC, wo EA (2 Questions)

Standard: D1D Ped Med Pt 1 Vignette D Risk Given: F, 8 9 11, A&GTC, w EA (2 Questions)

Group: Patient D2 - Ped Med

Standard: D2A Ped Med Pt 2 Vignette A Risk Est: F, 5 8 11, FM, wo EA (4 Questions)

Standard: D2B Ped Med Pt 2 Vignette B Risk Est: F, 5 8 11, FM, w EA (3 Questions)

Branch: New Branch

If

If Here is the first patient.  Sex: femaleFirst seizure: 5 years oldLast seizure: 8 years old (Durat... Is Displayed

Standard: D2C Ped Med Pt 2 Vignette C Risk Given: F, 5 8 11, FM, wo EA (2 Questions)

Standard: D2D Ped Med Pt 2 Vignette D Risk Est: F, 5 8 11, FM, w EA (2 Questions)

Branch: New Branch

If

If In the past year, which best describes your epilepsy/seizure patients? Mostly pediatric (17 years of age or younger) patients Is Selected

And In the past year, how many of the patients that you are managing for epilepsy had undergone epile... All Is Selected

EndSurvey: Advanced

Branch: New Branch

If

If In the past year, how many of the patients that you are managing for epilepsy had undergone epile... Some Is Selected

Or In the past year, how many of the patients that you are managing for epilepsy had undergone epile... All Is Selected

Standard: E1: General Questions - Surgical Only (5 Questions)

Branch: New Branch

If

If In the past year, how many of the patients that you are managing for epilepsy had undergone epile... Some Is Selected

Or In the past year, how many of the patients that you are managing for epilepsy had undergone epile... None Is Selected

Standard: E2: General Questions - Non-Surgical Only (8 Questions)

Standard: E3: General Questions - All (19 Questions)

Standard: F: Background (5 Questions)

EmbeddedData

AEDGroupValue will be set from Panel or URL.

age_truncValue will be set from Panel or URL.

COUNTRYValue will be set from Panel or URL.

GENDERValue will be set from Panel or URL.

PrimarySubspecialtyValue will be set from Panel or URL.

STATEValue will be set from Panel or URL.

UM_IDValue will be set from Panel or URL.

EndSurvey:

| Page Break |
| --- |

Start of Block: A: Screening Questions

| 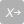 | 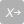 |
| --- | --- |

screen_US Prior to starting the survey, please use the link below to open and read the University of Michigan's informed consent form. Please keep a copy of this form for your records.

 [University of Michigan Informed Consent Form](https://aan.co1.qualtrics.com/WRQualtricsControlPanel/File.php?F=F_0B4kP7vy9nYbOXI)

 Are you willing to participate in this research?

- Yes (1)
- No (2)

Skip To: End of Survey If screen_US = 2

| Page Break |
| --- |

screen_infoA Thank you for agreeing to take our survey. The survey will take approximately 15 minutes to complete. The study has three parts:	Screening and eligibility.	Patient vignettes where we ask you to estimate future seizure risk and to provide your recommendation regarding whether the patient should withdraw their antiseizure medication now.	General questions pertaining to how you approach discontinuation and what would help you make decisions in the future.

| Page Break |
| --- |

screen_infoB First, we will ask you some questions to determine if you are eligible to take this survey.

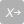

screen_neuro Are you a neurologist?

- Yes (1)
- No (2)

Skip To: End of Block If screen_neuro = 2

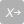

screen_practice In the past year, which describes your practice setting? *Mark all that apply.*

- Academic-based (1)
- Government-based (2)
- Community clinic-based (e.g., private practice, neurology, or multispecialty group) (3)
- ⊗None of the above, please describe (4) ________________________________________________
- ⊗I do not have a clinical practice (5)

Skip To: End of Block If screen_practice = 5

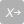

screen_seizure In the past year, how many of your patients were seen for seizure evaluation/management?

- None (1)
- Less than half (2)
- About half (3)
- More than half (4)

Skip To: End of Block If screen_seizure = 1

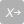

screen_surgery
In the past year, how many of the patients that you are managing for epilepsy had undergone epilepsy surgery?

- None (1)
- Some (2)
- All (3)

| Page Break |
| --- |

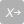

screen_advsped In the past year, which best describes your epilepsy/seizure patients?

- Mostly pediatric (17 years of age or younger) patients (1)
- Mostly adult (18 years of age or older) patients (2)

| Page Break |
| --- |

End of Block: A: Screening Questions

Start of Block: B1A Ad Med Pt 1 Vignette A: Risk Est, M, 26 28 30, F&BTC, wo EA

introAdMedA You are eligible, let's begin.

Although you might be familiar that a risk prediction calculator exists, we ask you not to use any outside resources as we are interested in how clinicians such as yourself intuitively estimate seizure risk.

| Page Break |
| --- |

patient01 Here is the first patient.
 **Sex:** male
 **First seizure:** 26 years old 
 **Last seizure:** 28 years old (Duration of seizures: 2 years)
 **Current age:** 30 years old (Duration seizure-free: 2 years)

 **Number of seizures:** 6
 **Semiology:** focal to bilateral tonic-clonic
 **Family history of seizures:** no
 **Febrile seizures:** yes
 **Developmentally:** normal
 **MRI:** nonlesional

 **Antiseizure medication:** levetiracetam
 **Today's EEG:** no epileptiform abnormalities

guess01 Estimate the risk of this patient having another seizure in the next two years if levetiracetam...

|  | % Risk |
| --- | --- |

|  | 0 | 10 | 20 | 30 | 40 | 50 | 60 | 70 | 80 | 90 | 100 |
| --- | --- | --- | --- | --- | --- | --- | --- | --- | --- | --- | --- |

| ... is continued () | 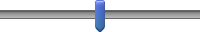 |
| --- | --- |
| ... withdrawal is started now () | 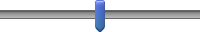 |

advise01_no How likely would you be to advise levetiracetam withdrawal if the patient has not experienced side-effects and if their job...

|  | Extremely  unlikely | Neither likely  nor unlikely | Extremely  likely |
| --- | --- | --- | --- |

|  | 0 | 1 | 2 | 3 | 4 | 5 | 6 | 7 | 8 | 9 | 10 |
| --- | --- | --- | --- | --- | --- | --- | --- | --- | --- | --- | --- |

| ... does require driving () | 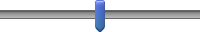 |
| --- | --- |
| ... does not require driving () | 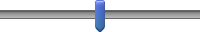 |

advise01_yes How likely would you be to advise levetiracetam withdrawal if the patient has experienced side-effects (bothersome mood changes and irritability) and if their job...

|  | Extremely  unlikely | Neither likely  nor unlikely | Extremely  likely |
| --- | --- | --- | --- |

|  | 0 | 1 | 2 | 3 | 4 | 5 | 6 | 7 | 8 | 9 | 10 |
| --- | --- | --- | --- | --- | --- | --- | --- | --- | --- | --- | --- |

| ... does require driving () | 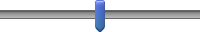 |
| --- | --- |
| ... does not require driving () | 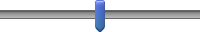 |

End of Block: B1A Ad Med Pt 1 Vignette A: Risk Est, M, 26 28 30, F&BTC, wo EA

Start of Block: B1B Ad Med Pt 1 Vignette B: Risk Est, M, 26 28 30, F&BTC, w EA

patient02 Now instead assume the patient’s current EEG shows **epileptiform abnormalities**.

 **Sex:** male
 **First seizure:** 26 years old
 **Last seizure:** 28 years old (Duration of seizures: 2 years)
 **Current age:** 30 years old (Duration seizure-free: 2 years)

 **Number of seizures:** 6
 **Semiology:** focal to bilateral tonic-clonic
 **Family history of seizures:** no
 **Febrile seizures:** yes
 **Developmentally:** normal
 **MRI:** nonlesional

 **Antiseizure medication:** levetiracetam
 **Today's EEG:** epileptiform abnormalities

guess02 Estimate the risk of this patient having another seizure in the next two years if levetiracetam ...

|  | % Risk |
| --- | --- |

|  | 0 | 10 | 20 | 30 | 40 | 50 | 60 | 70 | 80 | 90 | 100 |
| --- | --- | --- | --- | --- | --- | --- | --- | --- | --- | --- | --- |

| ... is continued () | 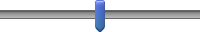 |
| --- | --- |
| ... withdrawal is started now () | 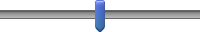 |

advise02_no How likely would you be to advise levetiracetam withdrawal if the patient has not experienced side-effects and if their job...

|  | Extremely  unlikely | Neither likely  nor unlikely | Extremely  likely |
| --- | --- | --- | --- |

|  | 0 | 1 | 2 | 3 | 4 | 5 | 6 | 7 | 8 | 9 | 10 |
| --- | --- | --- | --- | --- | --- | --- | --- | --- | --- | --- | --- |

| ... does require driving () | 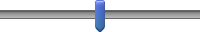 |
| --- | --- |

advise02_yes How likely would you be to advise levetiracetam withdrawal if the patient has experienced side-effects (bothersome mood changes and irritability) and if their job...

|  | Extremely  unlikely | Neither likely  nor unlikely | Extremely  likely |
| --- | --- | --- | --- |

|  | 0 | 1 | 2 | 3 | 4 | 5 | 6 | 7 | 8 | 9 | 10 |
| --- | --- | --- | --- | --- | --- | --- | --- | --- | --- | --- | --- |

| ... does not require driving () | 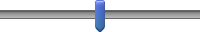 |
| --- | --- |

End of Block: B1B Ad Med Pt 1 Vignette B: Risk Est, M, 26 28 30, F&BTC, w EA

Start of Block: B1C Ad Med Pt 1 Vignette C: Risk Given, M, 26 28 30, F&BTC, wo EA

patient03
Next, we will show you similar questions, except we will include two hypothetical seizure probabilities.  First, assume the patient's current EEG shows **no epileptiform abnormalities**. 
 

The probability of another seizure in the next 2 years is:
     31% if levetiracetam is continued
     62% if levetiracetam withdrawal is started now

 **Sex:** male
 **First seizure:** 26 years old 
**Last seizure:** 28 years old (Duration of seizures: 2 years)
**Current age:** 30 years old (Duration seizure-free: 2 years)

 **Number of seizures:** 6
 **Semiology:** focal to bilateral tonic-clonic
 **Family history of seizures:** no
 **Febrile seizures:** yes
 **Developmentally:** normal
 **MRI:** nonlesional

 **Antiseizure medication:** levetiracetam
 **Today's EEG:** no epileptiform abnormalities

advise03_no How likely would you be to advise levetiracetam withdrawal if the patient has not experienced side-effects and if their job...

|  | Extremely  unlikely | Neither likely  nor unlikely | Extremely  likely |
| --- | --- | --- | --- |

|  | 0 | 1 | 2 | 3 | 4 | 5 | 6 | 7 | 8 | 9 | 10 |
| --- | --- | --- | --- | --- | --- | --- | --- | --- | --- | --- | --- |

| ... does require driving () | 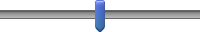 |
| --- | --- |

advise03_yes How likely would you be to advise levetiracetam withdrawal if the patient has experienced side-effects (bothersome mood changes and irritability) and if their job...

|  | Extremely  unlikely | Neither likely  nor unlikely | Extremely  likely |
| --- | --- | --- | --- |

|  | 0 | 1 | 2 | 3 | 4 | 5 | 6 | 7 | 8 | 9 | 10 |
| --- | --- | --- | --- | --- | --- | --- | --- | --- | --- | --- | --- |

| ... does not require driving () | 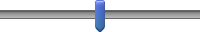 |
| --- | --- |

End of Block: B1C Ad Med Pt 1 Vignette C: Risk Given, M, 26 28 30, F&BTC, wo EA

Start of Block: B1D Ad Med Pt 1 Vignette D: Risk Given: M, 26 28 30, F&BTC, w EA

patient04 Next, the patient’s current EEG shows **epileptiform abnormalities**.  
 
The probability of another seizure in the next 2 years is:
      39% if levetiracetam is continue
       77% if levetiracetam withdrawal is started now 

 **Sex:** male
 **First seizure:** 26 years old 
**Last seizure:** 28 years old (Duration of seizures: 2 years)
**Current age:** 30 years old (Duration seizure-free: 2 years)

 **Number of seizures:** 6
 **Semiology:** focal to bilateral tonic-clonic
 **Family history of seizures:** no
 **Febrile seizures:** yes
 **Developmentally:** normal
 **MRI:** nonlesional

 **Antiseizure medication:** levetiracetam
 **Today's EEG:** epileptiform abnormalities

advise04_no How likely would you be to advise levetiracetam withdrawal if the patient has not experienced side-effects and if their job...

|  | Extremely  unlikely | Neither likely  nor unlikely | Extremely  likely |
| --- | --- | --- | --- |

|  | 0 | 1 | 2 | 3 | 4 | 5 | 6 | 7 | 8 | 9 | 10 |
| --- | --- | --- | --- | --- | --- | --- | --- | --- | --- | --- | --- |

| ... does require driving () | 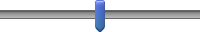 |
| --- | --- |

advise04_yes How likely would you be to advise levetiracetam withdrawal if the patient has experienced side-effects (bothersome mood changes and irritability) and if their job...

|  | Extremely  unlikely | Neither likely  nor unlikely | Extremely  likely |
| --- | --- | --- | --- |

|  | 0 | 1 | 2 | 3 | 4 | 5 | 6 | 7 | 8 | 9 | 10 |
| --- | --- | --- | --- | --- | --- | --- | --- | --- | --- | --- | --- |

| ... does not require driving () | 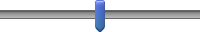 |
| --- | --- |

End of Block: B1D Ad Med Pt 1 Vignette D: Risk Given: M, 26 28 30, F&BTC, w EA

Start of Block: B2A Ad Med Pt 2 Vignette A: Risk Est, M, 26 28 38, F&BTC, wo EA

IntroAdMedB You are eligible, let's begin.

Although you might be familiar that a risk prediction calculator exists, we ask you not to use any outside resources as we are interested in how clinicians such as yourself intuitively estimate seizure risk.

| Page Break |
| --- |

patient05 Here is the first patient. 

 **Sex:** male
 **First seizure:** 26 years old
 **Last seizure:** 28 years old (Duration of seizures: 2 years)
 **Current age:** 38 years old (Duration seizure-free: 10 years)

 **Number of seizures:** 6
 **Semiology:** focal to bilateral tonic-clonic
 **Family history of seizures:** no
 **Febrile seizures:** yes
 **Developmentally:** normal
 **MRI:** nonlesional

 **Antiseizure medication:** levetiracetam
 **Today's EEG:** no epileptiform abnormalities

guess05 Estimate the risk of this patient having another seizure in the next two years if levetiracetam...

|  | % Risk |
| --- | --- |

|  | 0 | 10 | 20 | 30 | 40 | 50 | 60 | 70 | 80 | 90 | 100 |
| --- | --- | --- | --- | --- | --- | --- | --- | --- | --- | --- | --- |

| ... is continued () | 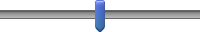 |
| --- | --- |
| ... withdrawal is started now () | 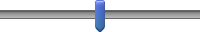 |

advise05_no How likely would you be to advise levetiracetam withdrawal if the patient has not experienced side-effects and if their job...

|  | Extremely  unlikely | Neither likely  nor unlikely | Extremely  likely |
| --- | --- | --- | --- |

|  | 0 | 1 | 2 | 3 | 4 | 5 | 6 | 7 | 8 | 9 | 10 |
| --- | --- | --- | --- | --- | --- | --- | --- | --- | --- | --- | --- |

| ... does require driving () | 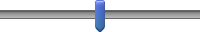 |
| --- | --- |
| ... does not require driving () | 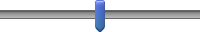 |

advise05_yes How likely would you be to advise levetiracetam withdrawal if the patient has experienced side-effects (bothersome mood changes and irritability) and if their job...

|  | Extremely  unlikely | Neither likely  nor unlikely | Extremely  likely |
| --- | --- | --- | --- |

|  | 0 | 1 | 2 | 3 | 4 | 5 | 6 | 7 | 8 | 9 | 10 |
| --- | --- | --- | --- | --- | --- | --- | --- | --- | --- | --- | --- |

| ... does require driving () | 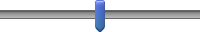 |
| --- | --- |
| ... does not require driving () | 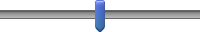 |

End of Block: B2A Ad Med Pt 2 Vignette A: Risk Est, M, 26 28 38, F&BTC, wo EA

Start of Block: B2B Ad Med Pt 2 Vignette B: Risk Est, M, 26 28 38, F&BTC, w EA

patient06
Now instead assume the patient's current EEG shows **epileptiform abnormalities**.
 Sex: male
 First seizure: 26 years old
 Last seizure: 28 years old (Duration of seizures: 2 years)
 Current age: 38 years old (Duration seizure-free: 10 years)
  
**Number of seizures:** 6
 **Semiology:** focal to bilateral tonic-clonic
 **Family history of seizures:** no
 **Febrile seizures:** yes
 **Developmentally:** normal
 **MRI:** nonlesional

 **Antiseizure medication:** levetiracetam
 **Today's EEG:** epileptiform abnormalities

guess06 Estimate the risk of this patient having another seizure in the next two years if levetiracetam ...

|  | % Risk |
| --- | --- |

|  | 0 | 10 | 20 | 30 | 40 | 50 | 60 | 70 | 80 | 90 | 100 |
| --- | --- | --- | --- | --- | --- | --- | --- | --- | --- | --- | --- |

| ... is continued () | 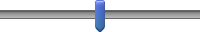 |
| --- | --- |
| ... withdrawal is started now () | 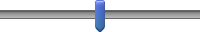 |

advise06_no How likely would you be to advise levetiracetam withdrawal if the patient has not experienced side-effects and if their job...

|  | Extremely  unlikely | Neither likely  nor unlikely | Extremely  likely |
| --- | --- | --- | --- |

|  | 0 | 1 | 2 | 3 | 4 | 5 | 6 | 7 | 8 | 9 | 10 |
| --- | --- | --- | --- | --- | --- | --- | --- | --- | --- | --- | --- |

| ... does require driving () | 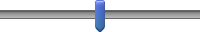 |
| --- | --- |

advise06_yes How likely would you be to advise levetiracetam withdrawal if the patient has experienced side-effects (bothersome mood changes and irritability) and if their job...

|  | Extremely  unlikely | Neither likely  nor unlikely | Extremely  likely |
| --- | --- | --- | --- |

|  | 0 | 1 | 2 | 3 | 4 | 5 | 6 | 7 | 8 | 9 | 10 |
| --- | --- | --- | --- | --- | --- | --- | --- | --- | --- | --- | --- |

| ... does not require driving () | 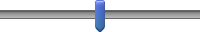 |
| --- | --- |

End of Block: B2B Ad Med Pt 2 Vignette B: Risk Est, M, 26 28 38, F&BTC, w EA

Start of Block: B2C Ad Med Pt 2 Vignette C: Risk Given, M, 26 28 38, F&BTC, wo EA

patient07

Next, we will show you similar questions, except we will include two hypothetical seizure probabilities.  First, assume the patient's current EEG shows **no epileptiform abnormalities**. 
 

The probability of another seizure in the next 2 years is:
     20% if levetiracetam is continued      40% if levetiracetam withdrawal is started now

 Sex: male
 First seizure: 26 years old
 Last seizure: 28 years old (Duration of seizures: 2 years)
Current age: 38 years old (Duration seizure-free: 10 years)

 **Number of seizures:** 6
 **Semiology:** focal to bilateral tonic-clonic
 **Family history of seizures:** no
 **Febrile seizures:** yes
 **Developmentally:** normal
 **MRI:** nonlesional

 **Antiseizure medication:** levetiracetam
 **Today's EEG:** no epileptiform abnormalities

advise07_No How likely would you be to advise levetiracetam withdrawal if the patient has not experienced side-effects and if their job...

|  | Extremely  unlikely | Neither likely  nor unlikely | Extremely  likely |
| --- | --- | --- | --- |

|  | 0 | 1 | 2 | 3 | 4 | 5 | 6 | 7 | 8 | 9 | 10 |
| --- | --- | --- | --- | --- | --- | --- | --- | --- | --- | --- | --- |

| ... does require driving () | 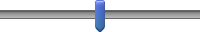 |
| --- | --- |

advise07_Yes How likely would you be to advise levetiracetam withdrawal if the patient has experienced side-effects (bothersome mood changes and irritability) and if their job...

|  | Extremely  unlikely | Neither likely  nor unlikely | Extremely  likely |
| --- | --- | --- | --- |

|  | 0 | 1 | 2 | 3 | 4 | 5 | 6 | 7 | 8 | 9 | 10 |
| --- | --- | --- | --- | --- | --- | --- | --- | --- | --- | --- | --- |

| ... does not require driving () | 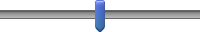 |
| --- | --- |

End of Block: B2C Ad Med Pt 2 Vignette C: Risk Given, M, 26 28 38, F&BTC, wo EA

Start of Block: B2D Ad Med Pt 2 Vignette D Risk Given: M, 26 28 38, F&BTC, w EA

patient08
Now instead assume the patient’s current EEG shows **epileptiform abnormalities**. 
 
The probability of another seizure in the next 2 years is:
      26% if levetiracetam is continued       52% if levetiracetam withdrawal is started now 

 Sex: male
 First seizure: 26 years old
 Last seizure: 28 years old (Duration of seizures: 2 years)
 Current age: 38 years old (Duration seizure-free: 10 years)
  
**Number of seizures:** 6
 **Semiology:** focal to bilateral tonic-clonic
 **Family history of seizures:** no
 **Febrile seizures:** yes
 **Developmentally:** normal
 **MRI:** nonlesional

 **Antiseizure medication:** levetiracetam
 **Today's EEG:** epileptiform abnormalities

advise08_no How likely would you be to advise levetiracetam withdrawal if the patient has not experienced side-effects and if their job...

|  | Extremely  unlikely | Neither likely  nor unlikely | Extremely  likely |
| --- | --- | --- | --- |

|  | 0 | 1 | 2 | 3 | 4 | 5 | 6 | 7 | 8 | 9 | 10 |
| --- | --- | --- | --- | --- | --- | --- | --- | --- | --- | --- | --- |

| ... does require driving () | 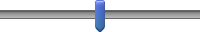 |
| --- | --- |

advise08_yes How likely would you be to advise levetiracetam withdrawal if the patient has experienced side-effects (bothersome mood changes and irritability) and if their job...

|  | Extremely  unlikely | Neither likely  nor unlikely | Extremely  likely |
| --- | --- | --- | --- |

|  | 0 | 1 | 2 | 3 | 4 | 5 | 6 | 7 | 8 | 9 | 10 |
| --- | --- | --- | --- | --- | --- | --- | --- | --- | --- | --- | --- |

| ... does not require driving () | 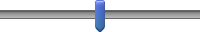 |
| --- | --- |

End of Block: B2D Ad Med Pt 2 Vignette D Risk Given: M, 26 28 38, F&BTC, w EA

Start of Block: C1A Ad Surg Pt 1 Vignette A: M, 30, F&BTC, 6M, wo EA

Display This Question:

If screen_surgery = 3

IntroAdSurgA1 You are eligible, let's begin.

Although you might be familiar that a risk prediction calculator exists, we ask you not to use any outside resources as we are interested in how clinicians such as yourself intuitively estimate seizure risk.

Display This Question:

If screen_surgery = 2

IntroAdSurgA2 Now we will show you some patients who have undergone epilepsy surgery. In this set, we will change both whether the EEG has epileptiform findings and also the seizure-free duration since surgery.

| Page Break |
| --- |

patient09 Here is the first patient.

 **Sex:** male
 **First seizure:** 15 years old 
 **Last seizure:** 30 years old (Duration of seizures: 15 years)
 **Current age:** 30 years old (Duration seizure-free: 6 months, since surgery)
  
**Semiology:** focal to bilateral tonic-clonic
 **Family history of seizures:** no
 **Febrile seizures:** yes
 **Developmentally:** normal
 **MRI and pathology:** left-sided mesial temporal sclerosis
   Antiseizure medication: levetiracetam
**Operation:** standard left-sided anterior temporal lobectomy with amygdala-hippocampectomy
 Operation date: 6 months ago
  
**Today's EEG:** no epileptiform abnormalities

guess09 Estimate the risk of another seizure in the next two years if levetiracetam…

|  | % Risk |
| --- | --- |

|  | 0 | 10 | 20 | 30 | 40 | 50 | 60 | 70 | 80 | 90 | 100 |
| --- | --- | --- | --- | --- | --- | --- | --- | --- | --- | --- | --- |

| ... is continued () | 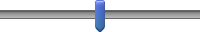 |
| --- | --- |
| ... withdrawal is started now () | 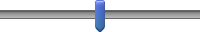 |

advise09_no How likely would you be to advise levetiracetam withdrawal if the patient has not experienced side-effects and if their job...

|  | Extremely  unlikely | Neither likely  nor unlikely | Extremely  likely |
| --- | --- | --- | --- |

|  | 0 | 1 | 2 | 3 | 4 | 5 | 6 | 7 | 8 | 9 | 10 |
| --- | --- | --- | --- | --- | --- | --- | --- | --- | --- | --- | --- |

| ...does require driving () | 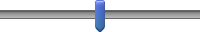 |
| --- | --- |
| ...does not require driving () | 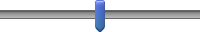 |

advise09_yes How likely would you be to advise levetiracetam withdrawal if the patient has experienced side-effects (bothersome mood changes and irritability) and if their job...

|  | Extremely  unlikely | Neither likely  nor unlikely | Extremely  likely |
| --- | --- | --- | --- |

|  | 0 | 1 | 2 | 3 | 4 | 5 | 6 | 7 | 8 | 9 | 10 |
| --- | --- | --- | --- | --- | --- | --- | --- | --- | --- | --- | --- |

| ...does require driving () | 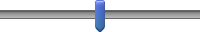 |
| --- | --- |
| ...does not require driving () | 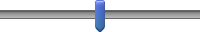 |

End of Block: C1A Ad Surg Pt 1 Vignette A: M, 30, F&BTC, 6M, wo EA

Start of Block: C1B Ad Surg Pt 1 Vignette B: M, 30, F&BTC, 6M, w EA

patient10 Now instead assume the patient’s current EEG shows **epileptiform abnormalities**.

 Sex: male
 First seizure: 15 (Duration of seizures: 15 years)
 Last seizure: 30
 Current age: 30 (Duration seizure-free: 6 months, since surgery)
  
Semiology: focal to bilateral tonic-clonic
 Family history of seizures: no
 Febrile seizures: yes
 Developmentally: normal
 MRI and pathology: left-sided mesial temporal sclerosis
   Antiseizure medication: levetiracetam
Operation: standard left-sided anterior temporal lobectomy with amygdala-hippocampectomy
 Operation date: 6 months ago
  
Today's EEG: epileptiform abnormalities

guess10 Estimate the risk of another seizure in the next two years if levetiracetam…

|  | % Risk |
| --- | --- |

|  | 0 | 10 | 20 | 30 | 40 | 50 | 60 | 70 | 80 | 90 | 100 |
| --- | --- | --- | --- | --- | --- | --- | --- | --- | --- | --- | --- |

| ... is continued () | 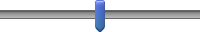 |
| --- | --- |
| ... withdrawal is started now () | 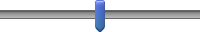 |

advise10_no How likely would you be to advise levetiracetam withdrawal if the patient has not experienced side-effects and if their job...

|  | Extremely  unlikely | Neither likely  nor unlikely | Extremely  likely |
| --- | --- | --- | --- |

|  | 0 | 1 | 2 | 3 | 4 | 5 | 6 | 7 | 8 | 9 | 10 |
| --- | --- | --- | --- | --- | --- | --- | --- | --- | --- | --- | --- |

| ...does require driving () | 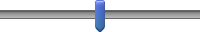 |
| --- | --- |

advise10_yes How likely would you be to advise levetiracetam withdrawal if the patient has experienced side-effects (bothersome mood changes and irritability) and if their job...

|  | Extremely  unlikely | Neither likely  nor unlikely | Extremely  likely |
| --- | --- | --- | --- |

|  | 0 | 1 | 2 | 3 | 4 | 5 | 6 | 7 | 8 | 9 | 10 |
| --- | --- | --- | --- | --- | --- | --- | --- | --- | --- | --- | --- |

| ...does not require driving () | 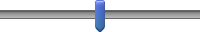 |
| --- | --- |

End of Block: C1B Ad Surg Pt 1 Vignette B: M, 30, F&BTC, 6M, w EA

Start of Block: C1C Ad Surg Pt 1 Vignette C: M, 30, F&BTC, 2Y, wo EA

patient11
Now instead assume **2 years seizure-free** and the patient’s current EEG shows **no epileptiform abnormalities.**  
Sex: male
 First seizure: 15
 Last seizure: 30 (Duration of seizures: 15 years)
 Current age: 32 (Duration seizure-free: 2 years, since surgery)
  
Semiology: focal to bilateral tonic-clonic
 Family history of seizures: no
 Febrile seizures: yes
 Developmentally: normal
 MRI and pathology: left-sided mesial temporal sclerosis
   Antiseizure medication: levetiracetam
Operation: standard left-sided anterior temporal lobectomy with amygdala-hippocampectomy
 Operation date: 2 years ago
  
Today's EEG: no epileptiform abnormalities

guess11 Estimate the risk of another seizure in the next two years if levetiracetam…

|  | % Risk |
| --- | --- |

|  | 0 | 10 | 20 | 30 | 40 | 50 | 60 | 70 | 80 | 90 | 100 |
| --- | --- | --- | --- | --- | --- | --- | --- | --- | --- | --- | --- |

| ... is continued () | 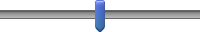 |
| --- | --- |
| ... withdrawal is started now () | 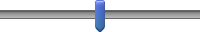 |

advise11_no How likely would you be to advise levetiracetam withdrawal if the patient has not experienced side-effects and if their job...

|  | Extremely  unlikely | Neither likely  nor unlikely | Extremely  likely |
| --- | --- | --- | --- |

|  | 0 | 1 | 2 | 3 | 4 | 5 | 6 | 7 | 8 | 9 | 10 |
| --- | --- | --- | --- | --- | --- | --- | --- | --- | --- | --- | --- |

| ...does require driving () | 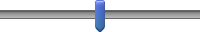 |
| --- | --- |

advise11_yes How likely would you be to advise levetiracetam withdrawal if the patient has experienced side-effects (bothersome mood changes and irritability) and if their job...

|  | Extremely  unlikely | Neither likely  nor unlikely | Extremely  likely |
| --- | --- | --- | --- |

|  | 0 | 1 | 2 | 3 | 4 | 5 | 6 | 7 | 8 | 9 | 10 |
| --- | --- | --- | --- | --- | --- | --- | --- | --- | --- | --- | --- |

| ...does not require driving () | 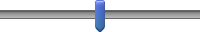 |
| --- | --- |

End of Block: C1C Ad Surg Pt 1 Vignette C: M, 30, F&BTC, 2Y, wo EA

Start of Block: C1D Ad Surg Pt 1 Vignette D: M, 30, F&BTC, 2Y, w EA

patient12 Now assume **epileptiform abnormalities**.
  
Sex: male
 First seizure: 15
 Last seizure: 30 (Duration of seizures: 15 years)
 Current age: 32 (Duration seizure-free: 2 years, since surgery)

 Semiology: focal to bilateral tonic-clonic
 Family history of seizures: no
 Febrile seizures: yes
 Developmentally: normal
 MRI and pathology: left-sided mesial temporal sclerosis

 Antiseizure medication: levetiracetam 
Operation: standard left-sided anterior temporal lobectomy with amygdala-hippocampectomy
 Operation date: 2 years ago

 Today's EEG: epileptiform abnormalities

guess12 Estimate the risk of another seizure in the next two years if levetiracetam…

|  | % Risk |
| --- | --- |

|  | 0 | 10 | 20 | 30 | 40 | 50 | 60 | 70 | 80 | 90 | 100 |
| --- | --- | --- | --- | --- | --- | --- | --- | --- | --- | --- | --- |

| ... is continued () | 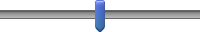 |
| --- | --- |
| ... withdrawal is started now () | 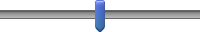 |

advise12_no How likely would you be to advise levetiracetam withdrawal if the patient has not experienced side-effects and if their job...

|  | Extremely  unlikely | Neither likely  nor unlikely | Extremely  likely |
| --- | --- | --- | --- |

|  | 0 | 1 | 2 | 3 | 4 | 5 | 6 | 7 | 8 | 9 | 10 |
| --- | --- | --- | --- | --- | --- | --- | --- | --- | --- | --- | --- |

| ...does require driving () | 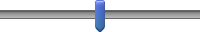 |
| --- | --- |

advise12_yes How likely would you be to advise levetiracetam withdrawal if the patient has experienced side-effects (bothersome mood changes and irritability) and if their job...

|  | Extremely  unlikely | Neither likely  nor unlikely | Extremely  likely |
| --- | --- | --- | --- |

|  | 0 | 1 | 2 | 3 | 4 | 5 | 6 | 7 | 8 | 9 | 10 |
| --- | --- | --- | --- | --- | --- | --- | --- | --- | --- | --- | --- |

| ...does not require driving () | 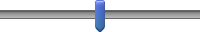 |
| --- | --- |

End of Block: C1D Ad Surg Pt 1 Vignette D: M, 30, F&BTC, 2Y, w EA

Start of Block: C2A Ad Surgery Pt 2 Vignette A: F, 30, F&BTC, 6M, wo EA

Display This Question:

If screen_surgery = 3

IntroAdSurgB1 You are eligible, let's begin.

Although you might be familiar that a risk prediction calculator exists, we ask you not to use any outside resources as we are interested in how clinicians such as yourself intuitively estimate seizure risk.

Display This Question:

If screen_surgery = 2

IntroAdSurgB2 Now we will show you some patients who have undergone epilepsy surgery. In this set, we will change both whether the EEG has epileptiform findings and also the seizure-free duration since surgery.

| Page Break |
| --- |

patient13 Here is the first patient. 

 **Sex:** female
 **First seizure:** 15
 **Last seizure:** 30 (Duration of seizures: 15 years)
 **Current age:** 30 (Duration seizure-free: 6 months, since surgery)

 **Semiology:** focal to bilateral tonic-clonic
 **Family history of seizures:** no
 **Febrile seizures:** no
 **Developmentally:** abnormal (mild intellectual deficits)
 **MRI and pathology:** focal cortical dysplasia type I
 Antiseizure medication: phenytoin

 **Operation:** complete resection of the right anterior temporal lobe and hippocampectomy
 **Operation date:** 6 months ago

 **Today’s EEG:** no epileptiform abnormalities

guess13 Estimate the risk of another seizure in the next two years if phenytoin...

|  | % Risk |
| --- | --- |

|  | 0 | 10 | 20 | 30 | 40 | 50 | 60 | 70 | 80 | 90 | 100 |
| --- | --- | --- | --- | --- | --- | --- | --- | --- | --- | --- | --- |

| ... is continued () | 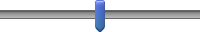 |
| --- | --- |
| ... withdrawal is started now () | 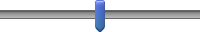 |

advise13_no How likely would you be to advise phenytoin withdrawal if the patient does not wish for pregnancy and if their job...

|  | Extremely  unlikely | Neither likely  nor unlikely | Extremely  likely |
| --- | --- | --- | --- |

|  | 0 | 1 | 2 | 3 | 4 | 5 | 6 | 7 | 8 | 9 | 10 |
| --- | --- | --- | --- | --- | --- | --- | --- | --- | --- | --- | --- |

| ...does require driving () | 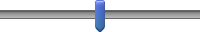 |
| --- | --- |
| ...does not require driving () | 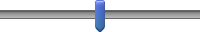 |

advise13_yes How likely would you be to advise phenytoin withdrawal if the patient does wish for pregnancy and if their job...

|  | Extremely  unlikely | Neither likely  nor unlikely | Extremely  likely |
| --- | --- | --- | --- |

|  | 0 | 1 | 2 | 3 | 4 | 5 | 6 | 7 | 8 | 9 | 10 |
| --- | --- | --- | --- | --- | --- | --- | --- | --- | --- | --- | --- |

| ...does require driving () | 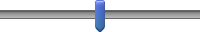 |
| --- | --- |
| ...does not require driving () | 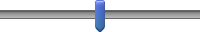 |

End of Block: C2A Ad Surgery Pt 2 Vignette A: F, 30, F&BTC, 6M, wo EA

Start of Block: C2B Ad Surgery Pt 2 Vignette B: F, 30, F&BTC, 6M, w EA

patient14 Now instead assume the patient’s current EEG shows **epileptiform abnormalities**.

 **Sex:** female
 **First seizure:** 15
 **Last seizure:** 30 (Duration of seizures: 15 years)
 **Current age:** 30 (Duration seizure-free: 6 months, since surgery)

 **Semiology:** focal to bilateral tonic-clonic
 **Family history of seizures:** no
 **Febrile seizures:** no
 **Developmentally:** abnormal (mild intellectual deficits)
 **MRI and pathology:** focal cortical dysplasia type I

 Antiseizure medication: phenytoin
 **Operation:** complete resection of the right anterior temporal lobe and hippocampectomy
 **Operation date:** 6 months ago

 **Today’s EEG:** epileptiform abnormalities

guess14 Estimate the risk of another seizure in the next two years if phenytoin...

|  | % Risk |
| --- | --- |

|  | 0 | 10 | 20 | 30 | 40 | 50 | 60 | 70 | 80 | 90 | 100 |
| --- | --- | --- | --- | --- | --- | --- | --- | --- | --- | --- | --- |

| ... is continued () | 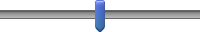 |
| --- | --- |
| ... withdrawal is started now () | 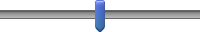 |

advise14_no How likely would you be to advise phenytoin withdrawal if the patient does not wish for pregnancy and if their job...

|  | Extremely  unlikely | Neither likely  nor unlikely | Extremely  likely |
| --- | --- | --- | --- |

|  | 0 | 1 | 2 | 3 | 4 | 5 | 6 | 7 | 8 | 9 | 10 |
| --- | --- | --- | --- | --- | --- | --- | --- | --- | --- | --- | --- |

| ...does require driving () | 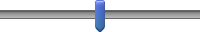 |
| --- | --- |

advise14_yes How likely would you be to advise phenytoin withdrawal if the patient does wish for pregnancy and if their job...

|  | Extremely  unlikely | Neither likely  nor unlikely | Extremely  likely |
| --- | --- | --- | --- |

|  | 0 | 1 | 2 | 3 | 4 | 5 | 6 | 7 | 8 | 9 | 10 |
| --- | --- | --- | --- | --- | --- | --- | --- | --- | --- | --- | --- |

| ...does not require driving () | 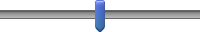 |
| --- | --- |

End of Block: C2B Ad Surgery Pt 2 Vignette B: F, 30, F&BTC, 6M, w EA

Start of Block: C2C Ad Surgery Pt 2 Vignette C: F, 30, F&BTC, 2Y, wo EA

patient15 Now assume **2 years seizure-free** and the patient's EEG shows **no epileptiform abnormalities**.

 Sex: female

 First seizure: 15
 Last seizure: 30 (Duration of seizures: 15 years)
 Current age: 32 (Duration seizure-free: 2 years, since surgery)

 Semiology: focal to bilateral tonic-clonic
 Family history of seizures: no
 Febrile seizures: no
 Developmentally: abnormal (mild intellectual deficits)
 MRI and pathology: focal cortical dysplasia type I

 Antiseizure medication: phenytoin
 Operation: complete resection of the right anterior temporal lobe and hippocampectomy
 Operation date: 2 years ago

 Today’s EEG: no epileptiform abnormalities

guess15 Estimate the risk of another seizure in the next two years if phenytoin...

|  | % Risk |
| --- | --- |

|  | 0 | 10 | 20 | 30 | 40 | 50 | 60 | 70 | 80 | 90 | 100 |
| --- | --- | --- | --- | --- | --- | --- | --- | --- | --- | --- | --- |

| ... is continued () | 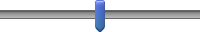 |
| --- | --- |
| ... withdrawal is started now () | 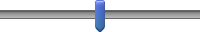 |

advise15_no How likely would you be to advise phenytoin withdrawal if the patient does not wish for pregnancy and if their job...

|  | Extremely  unlikely | Neither likely  nor unlikely | Extremely  likely |
| --- | --- | --- | --- |

|  | 0 | 1 | 2 | 3 | 4 | 5 | 6 | 7 | 8 | 9 | 10 |
| --- | --- | --- | --- | --- | --- | --- | --- | --- | --- | --- | --- |

| ...does require driving () | 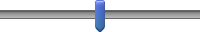 |
| --- | --- |

advise15_yes How likely would you be to advise levetiracetam withdrawal if the patient does wish for pregnancy and if their job...

|  | Extremely  unlikely | Neither likely  nor unlikely | Extremely  likely |
| --- | --- | --- | --- |

|  | 0 | 1 | 2 | 3 | 4 | 5 | 6 | 7 | 8 | 9 | 10 |
| --- | --- | --- | --- | --- | --- | --- | --- | --- | --- | --- | --- |

| ...does not require driving () | 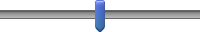 |
| --- | --- |

End of Block: C2C Ad Surgery Pt 2 Vignette C: F, 30, F&BTC, 2Y, wo EA

Start of Block: C2D Ad Surgery Pt 2 Vignette D: F, 30, F&BTC, 2Y, w EA

patient16 Now assume **epileptiform abnormalities**.

 Sex: female

 First seizure: 15
 Last seizure: 30 (Duration of seizures: 15 years)
 Current age: 32 (Duration seizure-free: 2 years, since surgery)

 Semiology: focal to bilateral tonic-clonic
 Family history of seizures: no
 Febrile seizures: no
 Developmentally: abnormal (mild intellectual deficits)
 MRI and pathology: focal cortical dysplasia type I

 Antiseizure medication: phenytoin
 Operation: complete resection of the right anterior temporal lobe and hippocampectomy
 Operation date: 2 years ago

 Today’s EEG:  epileptiform abnormalities

guess16 Estimate the risk of another seizure in the next two years if phenytoin...

|  | % Risk |
| --- | --- |

|  | 0 | 10 | 20 | 30 | 40 | 50 | 60 | 70 | 80 | 90 | 100 |
| --- | --- | --- | --- | --- | --- | --- | --- | --- | --- | --- | --- |

| ... is continued () | 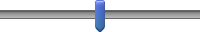 |
| --- | --- |
| ... withdrawal is started now () | 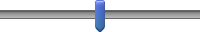 |

advise16_no How likely would you be to advise phenytoin withdrawal if the patient does not wish for pregnancy and if their job...

|  | Extremely  unlikely | Neither likely  nor unlikely | Extremely  likely |
| --- | --- | --- | --- |

|  | 0 | 1 | 2 | 3 | 4 | 5 | 6 | 7 | 8 | 9 | 10 |
| --- | --- | --- | --- | --- | --- | --- | --- | --- | --- | --- | --- |

| ...does require driving () | 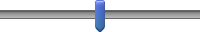 |
| --- | --- |

advise16_yes How likely would you be to advise levetiracetam withdrawal if the patient does wish for pregnancy and if their job...

|  | Extremely  unlikely | Neither likely  nor unlikely | Extremely  likely |
| --- | --- | --- | --- |

|  | 0 | 1 | 2 | 3 | 4 | 5 | 6 | 7 | 8 | 9 | 10 |
| --- | --- | --- | --- | --- | --- | --- | --- | --- | --- | --- | --- |

| ...does not require driving () | 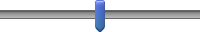 |
| --- | --- |

End of Block: C2D Ad Surgery Pt 2 Vignette D: F, 30, F&BTC, 2Y, w EA

Start of Block: D1A Ped Med Pt 1 Vignette A Risk Est: F, 8 9 11, A&GTC, wo EA

IntroPedMedA You are eligible, let's begin.

Although you might be familiar that a risk prediction calculator exists, we ask you not to use any outside resources as we are interested in how clinicians such as yourself intuitively estimate seizure risk.

| Page Break |
| --- |

patient17 Here is the first patient. 

**Sex:** female
 **First seizure:** 8
 **Last seizure:** 9 (Duration of seizures: 1 year)
 **Current age:** 11 (Duration seizure-free: 2 years)

 **Number of seizures:** countless absence
 **Semiology:** countless absence, 5 generalized tonic-clonic
 **Family history of seizures:** no
 **Febrile seizures:** no
 **Developmentally:** normal
 **MRI:** nonlesional

 **Antiseizure medication:** valproate
 **Today's EEG:** no epileptiform abnormalities

guess17 Estimate the risk of another seizure in the next two years if valproate...

|  | % Risk |
| --- | --- |

|  | 0 | 10 | 20 | 30 | 40 | 50 | 60 | 70 | 80 | 90 | 100 |
| --- | --- | --- | --- | --- | --- | --- | --- | --- | --- | --- | --- |

| ... is continued () | 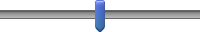 |
| --- | --- |
| ... withdrawal is started now () | 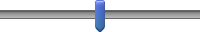 |

advise17 How likely would you be to advise valproate withdrawal if the patient...

|  | Extremely  unlikely | Neither | Extremely  likely |
| --- | --- | --- | --- |

|  | 0 | 1 | 2 | 3 | 4 | 5 | 6 | 7 | 8 | 9 | 10 |
| --- | --- | --- | --- | --- | --- | --- | --- | --- | --- | --- | --- |

| ... **has not experienced** side-effects () | 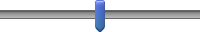 |
| --- | --- |
| ... **has experienced** side-effects (bothersome cognitive) () | 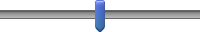 |

End of Block: D1A Ped Med Pt 1 Vignette A Risk Est: F, 8 9 11, A&GTC, wo EA

Start of Block: D1B Ped Med Pt 1 Vignette B Risk Est: F, 8 9 11, A&GTC, w EA

patient18 Now instead assume the patient’s current EEG shows **epileptiform abnormalities**.

 Sex: female
 First seizure: 8 years old
 Last seizure: 9 years old (Duration of seizures: 1 year)
 Current age: 11 years old (Duration seizure-free: 2 years)

 Number of seizures: countless absence
 Semiology: countless absence, 5 generalized tonic-clonic
 Family history of seizures: no
 Febrile seizures: no
 Developmentally: normal
 MRI: nonlesional

 Antiseizure medication: valproate
 Today's EEG: epileptiform abnormalities

guess18 Estimate the risk of another seizure in the next two years if valproate...

|  | % Risk |
| --- | --- |

|  | 0 | 10 | 20 | 30 | 40 | 50 | 60 | 70 | 80 | 90 | 100 |
| --- | --- | --- | --- | --- | --- | --- | --- | --- | --- | --- | --- |

| ... is continued () | 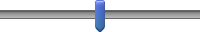 |
| --- | --- |
| ... withdrawal is started now () | 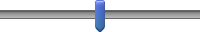 |

advise18 How likely would you be to advise valproate withdrawal if the patient...

|  | Extremely  unlikely | Neither | Extremely  likely |
| --- | --- | --- | --- |

|  | 0 | 1 | 2 | 3 | 4 | 5 | 6 | 7 | 8 | 9 | 10 |
| --- | --- | --- | --- | --- | --- | --- | --- | --- | --- | --- | --- |

| ... **has not experienced** side-effects () | 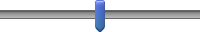 |
| --- | --- |
| ... **has experienced** side-effects (bothersome cognitive) () | 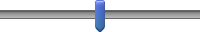 |

End of Block: D1B Ped Med Pt 1 Vignette B Risk Est: F, 8 9 11, A&GTC, w EA

Start of Block: D1C Ped Med Pt 1 Vignette C Risk Given: F, 8 9 11, A&GTC, wo EA

patient19

Next, we will show you similar questions, except we will include two hypothetical seizure probabilities.  First, assume the patient's current EEG shows **no epileptiform abnormalities**. 
 

The probability of another seizure in the next 2 years is:
     17% if valproate is continued
     34% if valproate withdrawal is started now
  Sex: female
 First seizure: 8 years old
 Last seizure: 9 years old (Duration of seizures: 1 year)
 Current age: 11 years old (Duration seizure-free: 2 years)

 Number of seizures: countless absence
 Semiology: countless absence, 5 generalized tonic-clonic
 Family history of seizures: no
 Febrile seizures: no
 Developmentally: normal
 MRI: nonlesional

 Antiseizure medication: valproate
 Today's EEG: no epileptiform abnormalities

advise19 How likely would you be to advise valproate withdrawal if the patient...

|  | Extremely  unlikely | Neither | Extremely  likely |
| --- | --- | --- | --- |

|  | 0 | 1 | 2 | 3 | 4 | 5 | 6 | 7 | 8 | 9 | 10 |
| --- | --- | --- | --- | --- | --- | --- | --- | --- | --- | --- | --- |

| ... **has not experienced** side-effects () | 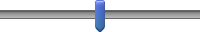 |
| --- | --- |
| ... **has experienced** side-effects (bothersome cognitive) () | 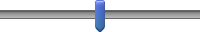 |

End of Block: D1C Ped Med Pt 1 Vignette C Risk Given: F, 8 9 11, A&GTC, wo EA

Start of Block: D1D Ped Med Pt 1 Vignette D Risk Given: F, 8 9 11, A&GTC, w EA

patient20
Next, the patient’s current EEG shows **epileptiform abnormalities**.  
 
The probability of another seizure in the next 2 years is:
      23% if valproate is continued
      46% if valproate withdrawal is started now 
  
Sex: female
First seizure: 8 years old
 Last seizure: 9 years old (Duration of seizures: 1 year)
Current age: 11 years old (Duration seizure-free: 2 years)

 Number of seizures: countless absence
 Semiology: countless absence, 5 generalized tonic-clonic
 Family history of seizures: no
 Febrile seizures: no
 Developmentally: normal
 MRI: nonlesional

 Antiseizure medication: valproate
 Today's EEG: epileptiform abnormalities

advise20 How likely would you be to advise valproate withdrawal if the patient...

|  | Extremely  unlikely | Neither | Extremely  likely |
| --- | --- | --- | --- |

|  | 0 | 1 | 2 | 3 | 4 | 5 | 6 | 7 | 8 | 9 | 10 |
| --- | --- | --- | --- | --- | --- | --- | --- | --- | --- | --- | --- |

| ... **has not experienced** side-effects () | 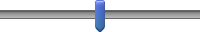 |
| --- | --- |
| ... **has experienced** side-effects (bothersome cognitive) () | 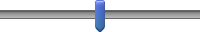 |

End of Block: D1D Ped Med Pt 1 Vignette D Risk Given: F, 8 9 11, A&GTC, w EA

Start of Block: D2A Ped Med Pt 2 Vignette A Risk Est: F, 5 8 11, FM, wo EA

IntroPedMedB You are eligible, let's begin.

Although you might be familiar that a risk prediction calculator exists, we ask you not to use any outside resources as we are interested in how clinicians such as yourself intuitively estimate seizure risk.

| Page Break |
| --- |

patient21
Here is the first patient. 

**Sex:** female
 **First seizure:** 5 years old
 **Last seizure:** 8 years old (Duration of seizures: 3 years)
 **Current age:** 11 years old (Duration seizure-free: 3 years)

 **Number of seizures:** 6
 **Semiology:** focal motor (left arm)
MRI and etiology: perinatal right-sided middle cerebral artery infarction
**Family history of seizures:** no
 **Febrile seizures:** yes
 **Developmentally:** moderate developmental delay

 **Antiseizure medication:** levetiracetam
 **Today's EEG:** no epileptiform abnormalities

guess21 Estimate the risk of another seizure in the next two years if levetiracetam...

|  | % Risk |
| --- | --- |

|  | 0 | 10 | 20 | 30 | 40 | 50 | 60 | 70 | 80 | 90 | 100 |
| --- | --- | --- | --- | --- | --- | --- | --- | --- | --- | --- | --- |

| ... is continued () | 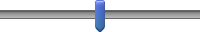 |
| --- | --- |
| ... withdrawal is started now () | 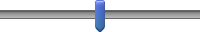 |

advise21 How likely would you be to advise levetiracetam withdrawal if the patient...

|  | Extremely  unlikely | Neither | Extremely  likely |
| --- | --- | --- | --- |

|  | 0 | 1 | 2 | 3 | 4 | 5 | 6 | 7 | 8 | 9 | 10 |
| --- | --- | --- | --- | --- | --- | --- | --- | --- | --- | --- | --- |

| ... **has not experienced** side-effects () | 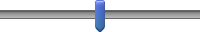 |
| --- | --- |
| ... **has experienced** side-effects (bothersome mood changes/irritability) () | 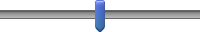 |

End of Block: D2A Ped Med Pt 2 Vignette A Risk Est: F, 5 8 11, FM, wo EA

Start of Block: D2B Ped Med Pt 2 Vignette B Risk Est: F, 5 8 11, FM, w EA

patient22
Now instead assume the patient’s current EEG shows **epileptiform abnormalities**.

 
Sex: female
 First seizure: 5 years old
 Last seizure: 8 years old (Duration of seizures: 3 years)
Current age: 11 years old (Duration seizure-free: 3 years)

 Number of seizures: 6
 Semiology: focal motor (left arm)
 MRI and etiology: perinatal right-sided middle cerebral artery infarction
Family history of seizures: no
 Febrile seizures: yes
 Developmentally: moderate developmental delay
  
Antiseizure medication: levetiracetam
 Today's EEG: epileptiform abnormalities

guess22 Estimate the risk of another seizure in the next two years if levetiracetam...

|  | % Risk |
| --- | --- |

|  | 0 | 10 | 20 | 30 | 40 | 50 | 60 | 70 | 80 | 90 | 100 |
| --- | --- | --- | --- | --- | --- | --- | --- | --- | --- | --- | --- |

| ... is continued () | 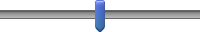 |
| --- | --- |
| ... withdrawal is started now () | 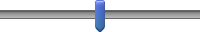 |

advise22 How likely would you be to advise levetiracetam withdrawal if the patient...

|  | Extremely  unlikely | Neither | Extremely  likely |
| --- | --- | --- | --- |

|  | 0 | 1 | 2 | 3 | 4 | 5 | 6 | 7 | 8 | 9 | 10 |
| --- | --- | --- | --- | --- | --- | --- | --- | --- | --- | --- | --- |

| ... **has not experienced** side-effects () | 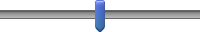 |
| --- | --- |
| ... **has experienced** side-effects (bothersome mood changes/irritability) () | 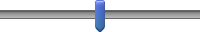 |

End of Block: D2B Ped Med Pt 2 Vignette B Risk Est: F, 5 8 11, FM, w EA

Start of Block: D2C Ped Med Pt 2 Vignette C Risk Given: F, 5 8 11, FM, wo EA

patient23

Next, we will show you similar questions, except we will include two hypothetical seizure probabilities.  First, assume the patient's current EEG shows **no epileptiform abnormalities**. 
 
**The probability of another seizure in the next 2 years is:**
     27% if levetiracetam is continued
      53% if levetiracetam withdrawal is started now
 

Sex: female
 First seizure: 5 years old
 Last seizure: 8 years old (Duration of seizures: 3 years)
Current age: 11 years old (Duration seizure-free: 3 years)

 Number of seizures: 6
 Semiology: focal motor (left arm)
 MRI and etiology: perinatal right-sided middle cerebral artery infarction
Family history of seizures: no
 Febrile seizures: yes
 Developmentally: moderate developmental delay
  
Antiseizure medication: levetiracetam
 Today's EEG: no epileptiform abnormalities

advise23 How likely would you be to advise levetiracetam withdrawal if the patient...

|  | Extremely  unlikely | Neither | Extremely  likely |
| --- | --- | --- | --- |

|  | 0 | 1 | 2 | 3 | 4 | 5 | 6 | 7 | 8 | 9 | 10 |
| --- | --- | --- | --- | --- | --- | --- | --- | --- | --- | --- | --- |

| ... **has not experienced** side-effects () | 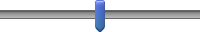 |
| --- | --- |
| ... **has experienced** side-effects (bothersome mood changes/irritability) () | 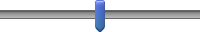 |

End of Block: D2C Ped Med Pt 2 Vignette C Risk Given: F, 5 8 11, FM, wo EA

Start of Block: D2D Ped Med Pt 2 Vignette D Risk Est: F, 5 8 11, FM, w EA

patient24
Next, the patient’s current EEG shows **epileptiform abnormalities**.  
 
**The probability of another seizure in the next 2 years is:**
       34% if levetiracetam is continued
      68% if levetiracetam withdrawal is started now 
  

Sex: female
 First seizure: 5 years old
 Last seizure: 8 years old (Duration of seizures: 3 years)
Current age: 11 years old (Duration seizure-free: 3 years)

 Number of seizures: 6
 Semiology: focal motor (left arm)
 MRI and etiology: perinatal right-sided middle cerebral artery infarction
Family history of seizures: no
 Febrile seizures: yes
 Developmentally: moderate developmental delay
  
Antiseizure medication: levetiracetam
 Today's EEG: epileptiform abnormalities

advise24 How likely would you be to advise levetiracetam withdrawal if the patient...

|  | Extremely  unlikely | Neither | Extremely  likely |
| --- | --- | --- | --- |

|  | 0 | 1 | 2 | 3 | 4 | 5 | 6 | 7 | 8 | 9 | 10 |
| --- | --- | --- | --- | --- | --- | --- | --- | --- | --- | --- | --- |

| ... **has not experienced** side-effects () | 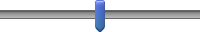 |
| --- | --- |
| ... **has experienced** side-effects (bothersome mood changes/irritability) () | 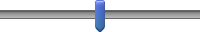 |

End of Block: D2D Ped Med Pt 2 Vignette D Risk Est: F, 5 8 11, FM, w EA

Start of Block: E1: General Questions - Surgical Only

gensurg_intro The following are general questions on treatment decisions for surgical patients, not specific to any single vignette.

gensurg_monopolyyes After anticipated curative surgery if a patient has been seizure-free for awhile, do you ever advise the withdrawal of antiseizure medication(s) from a patient who is currently on...

|  | Yes (1) | No, I don't advise stopping (2) |
| --- | --- | --- |
| 1 antiseizure med. (1) |  |  |
| >1 antiseizure med. (2) |  |  |

| Page Break |
| --- |

Display This Question:

If gensurg_monopolyyes = 1 [ 1 ]

Or gensurg_monopolyyes = 2 [ 1 ]

gensurg_startwd For patients after anticipated curative surgery: 

How many years of seizure-freedom do you typically require before advising to start withdrawal if the patient is on...

Display This Choice:

If gensurg_monopolyyes = 1 [ 1 ]

- 1 antiseizure med. (1) ________________________________________________

Display This Choice:

If gensurg_monopolyyes = 2 [ 1 ]

- >1 antiseizure med. (4) ________________________________________________

Display This Question:

If gensurg_monopolyyes = 1 [ 1 ]

Or gensurg_monopolyyes = 2 [ 1 ]

genmed_stop Over how many months do you typically suggest tapering until completely stopping antiseizure medication if the patient is on...

Display This Choice:

If gensurg_monopolyyes = 1 [ 1 ]

- 1 antiseizure med. (1) ________________________________________________

Display This Choice:

If gensurg_monopolyyes = 2 [ 1 ]

- >1 antiseizure med. (4) ________________________________________________

| Page Break |
| --- |

gensurg_relapserisk A patient underwent anticipated curative surgery.  Below what seizure relapse risk (within the next 2 years) would you advise starting withdrawal if the patient has...

|  | % Risk |
| --- | --- |

|  | 0 | 10 | 20 | 30 | 40 | 50 | 60 | 70 | 80 | 90 | 100 |
| --- | --- | --- | --- | --- | --- | --- | --- | --- | --- | --- | --- |

| ... convulsive seizures? () | 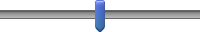 |
| --- | --- |
| ... nonconvulsive seizures? () | 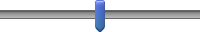 |

| Page Break |
| --- |

End of Block: E1: General Questions - Surgical Only

Start of Block: F: Background

demo_intro Now we will just ask you a few questions about yourself, and then the survey will be done in a moment.

| Page Break |
| --- |

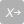

demo_boardUS Are you board-certified in epilepsy or clinical neurophysiology? *Mark all that apply.*

- Epilepsy (1)
- Clinical neurophysiology (2)
- ⊗Neither epilepsy nor clinical neurophysiology (3)

| Page Break |
| --- |

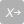

demo_yrspractice How many years have you been treating patients with epilepsy?

- 0-5 years (1)
- 6-10 years (2)
- 11-15 years (3)
- 16-20 years (4)
- More than 20 years (5)

| Page Break |
| --- |

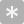

demo__percentclinic What percent of your job effort is clinical (0-100%)?

________________________________________________________________

| Page Break |
| --- |

anythingelse Please let us know anything else you wish about challenges you face with discontinuation decisions, and what would help you make these decisions?

________________________________________________________________

________________________________________________________________

________________________________________________________________

________________________________________________________________

________________________________________________________________

End of Block: F: Background
